# Development and validation of the 4C Deterioration model for adults hospitalised with COVID-19

**DOI:** 10.1101/2020.10.09.20209957

**Authors:** Rishi K Gupta, Ewen M Harrison, Antonia Ho, Annemarie B Docherty, Stephen R Knight, Maarten van Smeden, Ibrahim Abubakar, Marc Lipman, Matteo Quartagno, Riinu Pius, Iain Buchan, Gail Carson, Thomas M Drake, Jake Dunning, Cameron J Fairfield, Carrol Gamble, Christopher A Green, Sophie Halpin, Hayley E Hardwick, Karl A Holden, Peter W Horby, Clare Jackson, Kenneth A Mclean, Laura Merson, Jonathan S Nguyen-Van-Tam, Lisa Norman, Piero L Olliaro, Mark G Pritchard, Clark D Russell, James Scott-Brown, Catherine A Shaw, Aziz Sheikh, Tom Solomon, Cathie Sudlow, Olivia V Swann, Lance Turtle, Peter JM Openshaw, J Kenneth Baillie, Malcolm G Semple, Mahdad Noursadeghi, on behalf of the ISARIC4C Investigators

## Abstract

Prognostic models to predict the risk of clinical deterioration in acute COVID-19 are required to inform clinical management decisions. Among 75,016 consecutive adults across England, Scotland and Wales prospectively recruited to the ISARIC Coronavirus Clinical Characterisation Consortium (ISARIC4C) study, we developed and validated a multivariable logistic regression model for in-hospital clinical deterioration (defined as any requirement of ventilatory support or critical care, or death) using 11 routinely measured variables. We used internal-external cross-validation to show consistent measures of discrimination, calibration and clinical utility across eight geographical regions. We further validated the final model in held-out data from 8,252 individuals in London, with similarly consistent performance (C-statistic 0.77 (95% CI 0.75 to 0.78); calibration-in-the-large 0.01 (−0.04 to 0.06); calibration slope 0.96 (0.90 to 1.02)). Importantly, this model demonstrated higher net benefit than using other candidate scores to inform decision-making. Our 4C Deterioration model thus demonstrates unprecedented clinical utility and generalisability to predict clinical deterioration among adults hospitalised with COVID-19.

## Introduction

The coronavirus disease 2019 (COVID-19) pandemic continues to overwhelm healthcare systems worldwide^1^. Effective triage of patients presenting to hospital for risk of progressive deterioration is critical to inform clinical decision-making and facilitate effective resource allocation. Moreover, early identification of higher risk subgroups enables targeted recruitment for randomised controlled trials of therapies with equipoise^2^, and more precise delivery of treatments for which effectiveness is known to vary according to disease severity^3–5^.

A large number of multivariable clinical prognostic models for patients with COVID-19 have rapidly accrued to predict adverse outcomes of mortality or clinical deterioration^6^. The vast majority of candidate models subjected to comprehensive quality assessment have been classified as being at high risk of bias, and therefore may not be generalisable^6,7^. Moreover, none of the multivariable prognostic models included in a systematic head-to-head external validation study outperformed univariable predictors^8^, highlighting a critical need to adhere to rigorous model development methodology using large scale multi-site data, in order to facilitate generalisability.

We have previously reported a pragmatic prognostic index for in-hospital mortality from the ISARIC Coronavirus Clinical Characterisation Consortium (ISARIC4C) study^9^. Here, we extend this work through a larger study cohort to develop and validate a prognostic model for in-hospital clinical deterioration. We use the wide geographic coverage of the ISARIC4C study cohort in England, Wales and Scotland to explore between-region heterogeneity, and to comprehensively assess model generalisability with respect to discrimination, calibration and clinical utility. We have called this the 4C Deterioration model.

## Methods

### Study population and data collection

The International Severe Acute Respiratory and emerging Infections Consortium (ISARIC) World Health Organization (WHO) Clinical Characterisation Protocol UK (CCP-UK) study is being conducted by the ISARIC Coronavirus Clinical Characterisation Consortium (ISARIC4C) in 260 hospitals across England, Scotland, and Wales (National Institute for Health Research Clinical Research Network Central Portfolio Management System ID 14152). Further details of this prospective cohort study have been reported previously^10^. In this analysis, we included consecutive adults (≥18 years) with highly suspected or confirmed COVID-19 in whom eligibility criteria were confirmed. The study is reported in accordance with transparent reporting of a multivariable prediction model for individual prognosis or diagnosis (TRIPOD) guidance^11^. Demographic, clinical, and outcome data were collected through a standardised case record form as reported previously^9,10,12^. In this analysis, hospitals were classified and analysed by UK National Health Service (NHS) region^13^.

### Outcomes

We defined a composite primary outcome of in-hospital ‘clinical deterioration’ that includes: (a) initiation of ventilatory support (non-invasive ventilation, invasive mechanical ventilation or extra-corporeal membrane oxygenation); (b) high-dependency or intensive care unit admission; or (c) death. This composite outcome aligns closely with World Health Organization Clinical Progression Scale ≥6 and ensures that the outcome is generalisable between hospitals, since respiratory support practices may vary considerably^14^. We only included participants admitted or first assessed for COVID-19 until 26/08/2020 in order to allow a minimum four-week interval for registration of outcome events (until final data extraction date of 24/09/2020). Participants who had ongoing hospital care at the end of follow-up were classified as not meeting the endpoint, since the risk of deterioration declines with time since admission^8^. Missing outcomes were imputed, as described below.

### Candidate predictors

We specified candidate predictor variables *a priori* based on previous prognostic scores, and emerging literature describing routinely measured biomarkers associated with COVID-19 prognosis, as described previously^9^. We only considered predictors available in at least 60% of the study population for further analysis. In addition to the variables included in our previous analysis, we included nosocomial COVID-19 acquisition as an additional candidate predictor, since we hypothesised that acquisition of infection in hospital may be associated with differential outcomes. Community-acquired infection was defined as symptom onset or first positive SARS-CoV-2 PCR within 7 days from admission; participants who did not meet these criteria and had either symptom onset or first positive SARS-CoV-2 PCR >7 days from admission were classified as nosocomial cases^15^. Nosocomial cases who met the deterioration outcome prior to onset of COVID-19 were excluded.

Comorbidities were defined according to a modified Charlson comorbidity index^16^, with the addition of clinician-defined obesity, as reported previously^9^. We considered a composite ‘number of comorbidities’ variable for inclusion in the model, which included the following co-morbidities: chronic cardiac disease, chronic respiratory disease (excluding asthma), chronic renal disease, mild to severe liver disease, dementia, chronic neurological disease, connective tissue disease, diabetes mellitus, HIV or AIDS, malignancy and clinician-defined obesity.

All predictors were taken from the day of hospital admission, or the day of first clinical suspicion of COVID-19 for patients with nosocomial infection. Continuous predictors were modelled with restricted cubic splines using a default of four knots, placed at recommended locations based on percentiles^17^. Glasgow coma scale was categorised as 15 *vs*. <15 since there were insufficient data points below 15 to fit spline functions.

### Sample size

We assumed *a priori* that at least 30% of hospital admission would reach the primary outcome. The most comparable existing tools for risk stratification are the qSOFA, NEWS2, and CURB-65 scores^18–20^. qSOFA achieved C-statistics ranging from 0.71-0.78 during external validation^18^. Based on a conservative C-statistic of 0.71, we assumed a Cox-Snell R^2^ of 0.24. With 40 candidate predictor parameters (including transformations), the minimum sample size required for new model development was estimated to be 1,290, with 387 events^21,22^.

### Missing data

We handled missing data (including predictors and outcomes) using multiple imputation with chained equations, assuming missingness at random^23^, using the mice package in R^24^. We included all predictors (including restricted cubic spline transformations) and the outcome in the imputation models to ensure compatibility. Imputation was done separately for each NHS region to preserve potential inter-region heterogeneity. We generated 10 multiply imputed datasets; all primary analyses were performed in each imputed dataset and model and validation parameters were pooled using Rubin’s rules^25^.

### Model development

We hypothesised that heterogeneity among populations, healthcare services and clinical management may be present between NHS regions, which may contribute to differences in model performance; we therefore split the dataset according to the region of the contributing hospital. We included eight regions in model development and internal-external cross validation (East of England, the Midlands, North East England and Yorkshire, North West England, Scotland, South East England, South West England and Wales), with one region held out for further validation (London).

We used a logistic regression modelling approach and performed backward elimination of the *a priori* candidate variables using Akaike information criterion (AIC). This process was done separately in each multiply imputed dataset and in each NHS region in the development set. Predictors were required to be retained in >50% of multiply imputed datasets in >50% of development NHS regions in order to enter the final model. We specified this in order to retain a parsimonious set of predictors that had consistent prognostic value across the development NHS regions.

### Internal-external cross-validation

The model including the selected variables was then validated in the development dataset using the internal-external cross-validation framework, in order to concurrently examine between-region heterogeneity and assess generalisability^26^. During each internal-external cross-validation cycle, one of the contributing NHS regions within the development set was iteratively discarded from model training. Validation was then evaluated in the omitted NHS region by quantifying the model C-statistic, calibration slope and calibration-in-the-large^27^. We used random-effects meta-analysis to calculate pooled C-statistic, calibration slope and calibration-in-the-large statistics from internal-external cross-validation, and forest plots were examined to assess heterogeneity between regions. Calibration plots were also generated for each internal-external cross-validation cycle by fitting a loess smoother between the model predictions and the outcome in the stacked multiply imputed datasets. Recalibration to each region was performed by re-estimating the model intercept in the validation sets during each internal-external cross-validation cycle.

Decision curve analysis was also done in the internal-external cross-validation validation sets to quantify the net benefit of implementing the model in clinical practice^28^, compared to: (a) a ‘treat all’ approach; (b) a ‘treat none’ approach; and (c) using other candidate generic and COVID-specific clinical prognostic models to stratify treatment, identified by recent systematic reviews^6,8,9^. Only candidate models where constituent variables were available among >60% of the cohort were considered. Candidate models using points scores were calibrated to the validation data during decision curve analysis, resulting in optimistic estimates of their net benefit. All decision curves were loess-smoothed, from stacked multiply imputed datasets.

### Model validation in held-out NHS region

The final model was then trained using the full development dataset and validation was further evaluated in the held-out NHS region (London) by quantifying the C-statistic, calibration slope and calibration-in-the-large, and by visualisation of calibration plots^27^. Decision curve analysis was also performed, with comparison to other candidate models.

All analyses were conducted in R (version 3.6.3) using tidyverse (version 1.3.0)^29^, rms (version 6.0-1)^30^ for logistic regression modelling, mice (version 3.11.0)^24^ for multiple imputation and rmda (version 1.6)^31^ for decision curve analysis.

### Sensitivity analyses

We assessed validation of the final model using complete case data only in the held-out NHS region. We also recalculated validation metrics when: (a) excluding participants who experienced the outcome on the day of admission, in order to assess discrimination of the final model without these early events; (b) excluding participants in the validation cohort who had ongoing hospital care at the end of follow-up; (c) stratifying the validation cohort by community *vs*. nosocomial infection; and (d) excluding community-acquired cases who developed symptoms in the interval between admission and the temporal threshold for nosocomial infection, in order to assess any effect of incorrect inclusion of nosocomial infections within the community acquired cases. We also repeated the analysis using an alternative multiple imputation approach, using the aregImpute function from the rms package in R^30^, and recalculated model parameters using alternative temporal definitions of nosocomial SARS-CoV-2 infection (>5 days and >10 days after admission, compared to >7 days in the primary analysis).

### Ethical approval

Ethical approval was given by the South Central-Oxford C Research Ethics Committee in England (reference 13/SC/0149), and by the Scotland A Research Ethics Committee (reference 20/SS/0028). The study was registered at https://www.isrctn.com/ISRCTN66726260.

## Results

### Overview of study cohort

A total of 75,016 adults were recruited to the ISARIC4C study during the study period. Baseline demographic, physiological and laboratory characteristics of the cohort are shown stratified by outcome in Table 1 and by community *vs*. nosocomial infection in Supplementary Table 1. A total of 31,993/74,018 (43.2%) participants with outcomes available met the composite primary outcome of clinical deterioration during hospital admission. The interval between hospital admission and deterioration events, stratified by deterioration category, is shown in Supplementary Figure 1 (median time to deterioration 4 days; interquartile range 1-9). The overall risk of deterioration generally declined with increasing time from admission, supporting our approach to classify patients requiring ongoing hospital care at the end of follow-up as not meeting an endpoint in the primary analysis. Outcomes were missing for 998/75,016 (1.3%) participants.

**Table 1:** Baseline characteristics of the study cohort, stratified by outcome. Participants are shown by the first chronological deterioration category through which they met the composite primary outcome (high dependency unit (HDU) admission, intensive care (ICU) admission or ventilatory support, or death).

| Characteristic | Overall, N = 75,016 | Ventilatory support or HDU/ICU, N = 15,089<sup>1</sup> | Died, N = 16,904<sup>1</sup> | No deterioration, N = 42,025<sup>1</sup> | (Missing), N = 998<sup>1</sup> |
| --- | --- | --- | --- | --- | --- |
| Age (years) | 75 (60, 84) | 65 (55, 75) | 83 (77, 89) | 73 (57, 83) | 75 (59, 84) |
| Sex |  |  |  |  |  |
| Female | 32,846 (44%) | 5,156 (34%) | 7,114 (42%) | 20,142 (48%) | 434 (44%) |
| Male | 42,026 (56%) | 9,910 (66%) | 9,753 (58%) | 21,800 (52%) | 563 (56%) |
| Missing | 144 | 23 | 37 | 83 | 1 |
| Ethnicity |  |  |  |  |  |
| White | 55,079 (83%) | 9,985 (75%) | 13,628 (90%) | 30,855 (83%) | 611 (79%) |
| South Asian | 3,520 (5.3%) | 1,010 (7.6%) | 479 (3.1%) | 1,992 (5.4%) | 39 (5.1%) |
| Black | 2,554 (3.8%) | 744 (5.6%) | 345 (2.3%) | 1,435 (3.9%) | 30 (3.9%) |
| East Asian | 492 (0.7%) | 162 (1.2%) | 71 (0.5%) | 255 (0.7%) | 4 (0.5%) |
| Other | 4,848 (7.3%) | 1,406 (11%) | 699 (4.6%) | 2,656 (7.1%) | 87 (11%) |
| Missing | 8,523 | 1,782 | 1,682 | 4,832 | 227 |
| Number of comorbidities | 1 (1, 2) | 1 (0, 2) | 2 (1, 3) | 1 (0, 2) | 1 (0, 2) |
| Missing | 840 | 68 | 203 | 406 | 163 |
| Nosocomial infection | 7,385 (9.9%) | 592 (3.9%) | 2,111 (13%) | 4,538 (11%) | 144 (17%) |
| Missing | 636 | 62 | 101 | 345 | 128 |
| Radiographic infiltrates | 29,594 (62%) | 8,427 (77%) | 6,965 (63%) | 14,015 (55%) | 187 (54%) |
| Missing | 27,245 | 4,132 | 5,929 | 16,534 | 650 |
| Temperature (°C) | 37.2 (36.5, 38.1) | 37.5 (36.8, 38.4) | 37.1 (36.4, 38.0) | 37.1 (36.5, 38.0) | 37.0 (36.5, 38.0) |
| Missing | 3,110 | 463 | 650 | 1,783 | 214 |
| Heart rate (per min) | 90 (78, 104) | 95 (82, 109) | 90 (77, 105) | 88 (76, 102) | 89 (78, 102) |
| Missing | 3,389 | 424 | 721 | 2,021 | 223 |
| Respiratory rate (per min) | 20 (18, 26) | 24 (20, 30) | 22 (18, 28) | 20 (18, 24) | 20 (18, 24) |
| Missing | 3,537 | 483 | 689 | 2,113 | 252 |
| Systolic blood pressure (mmHg) | 130 (114, 147) | 129 (115, 145) | 128 (110, 147) | 130 (115, 147) | 130 (114, 148) |
| Missing | 3,192 | 428 | 651 | 1,891 | 222 |
| Diastolic blood pressure (mmHg) | 74 (64, 84) | 74 (65, 83) | 71 (61, 82) | 75 (65, 84) | 73 (65, 83) |
| Missing | 3,335 | 460 | 693 | 1,955 | 227 |
| SpO2 (%) | 95 (92, 97) | 94 (89, 96) | 95 (91, 97) | 96 (94, 97) | 96 (93, 98) |
| Missing | 3,760 | 538 | 802 | 2,203 | 217 |
| Room air or oxygen |  |  |  |  |  |
| Room air | 48,626 (69%) | 7,249 (50%) | 9,991 (63%) | 30,810 (79%) | 576 (76%) |
| Oxygen | 21,469 (31%) | 7,141 (50%) | 5,827 (37%) | 8,324 (21%) | 177 (24%) |
| Missing | 4,921 | 699 | 1,086 | 2,891 | 245 |
| Glasgow coma scale | 15 (15, 15) | 15 (15, 15) | 15 (15, 15) | 15 (15, 15) | 15 (15, 15) |
| Missing | 7,849 | 1,762 | 1,697 | 4,021 | 369 |
| Haemoglobin (g/L) | 128 (112, 142) | 132 (116, 145) | 122 (105, 138) | 129 (113, 142) | 128 (112, 141) |
| Missing | 11,781 | 1,422 | 2,568 | 7,449 | 342 |
| White cell count (x10<sup>9</sup>/L) | 7.5 (5.4, 10.7) | 8.0 (5.7, 11.2) | 8.3 (5.8, 11.9) | 7.1 (5.2, 9.9) | 7.7 (5.4, 10.7) |
| Missing | 12,163 | 1,515 | 2,675 | 7,624 | 349 |
| Lymphocytes (x10<sup>9</sup>/L) | 0.90 (0.60, 1.30) | 0.80 (0.60, 1.20) | 0.80 (0.50, 1.16) | 0.96 (0.67, 1.40) | 0.90 (0.60, 1.30) |
| Missing | 12,377 | 1,558 | 2,706 | 7,764 | 349 |
| Neutrophils (x10<sup>9</sup>/L) | 5.8 (3.9, 8.7) | 6.4 (4.3, 9.3) | 6.6 (4.3, 9.9) | 5.3 (3.6, 7.9) | 5.8 (3.7, 8.7) |
| Missing | 12,341 | 1,557 | 2,708 | 7,727 | 349 |
| Platelets (x10<sup>9</sup>/L) | 221 (167, 291) | 219 (166, 287) | 209 (154, 284) | 226 (173, 294) | 218 (161, 285) |
| Missing | 12,496 | 1,562 | 2,736 | 7,848 | 350 |
| Alanine aminotransferase (IU/L) | 25 (16, 43) | 33 (21, 54) | 22 (15, 37) | 24 (15, 40) | 24 (15, 43) |
| Missing | 26,783 | 3,895 | 6,300 | 16,086 | 502 |
| Bilirubin (mg/dL) | 10 (7, 14) | 10 (7, 15) | 10 (7, 15) | 9 (6, 13) | 10 (7, 14) |
| Missing | 22,979 | 3,143 | 5,317 | 14,026 | 493 |
| Urea (mmol/L) | 7 (5, 11) | 7 (5, 11) | 10 (7, 16) | 6 (4, 9) | 7 (5, 12) |
| Missing | 18,554 | 2,794 | 3,974 | 11,345 | 441 |
| Creatinine (μmol/L) | 86 (67, 121) | 86 (68, 118) | 106 (76, 158) | 81 (64, 107) | 85 (68, 122) |
| Missing | 12,693 | 1,636 | 2,706 | 7,999 | 352 |
| Sodium (mmol/L) | 137 (134, 140) | 136 (133, 139) | 138 (135, 143) | 137 (134, 140) | 137 (134, 140) |
| Missing | 12,312 | 1,515 | 2,631 | 7,811 | 355 |
| C-reactive protein (mg/L) | 80 (33, 154) | 126 (64, 210) | 98 (48, 174) | 58 (22, 119) | 76 (30, 151) |
| Missing | 16,353 | 2,223 | 3,515 | 10,203 | 412 |
| NHS region |  |  |  |  |  |
| East of England | 7,865 (10%) | 1,653 (11%) | 1,935 (11%) | 4,223 (10%) | 54 (5.4%) |
| London | 8,252 (11%) | 2,287 (15%) | 1,510 (8.9%) | 4,400 (10%) | 55 (5.5%) |
| Midlands | 15,599 (21%) | 2,559 (17%) | 3,703 (22%) | 9,068 (22%) | 269 (27%) |
| North East and Yorkshire | 10,310 (14%) | 2,236 (15%) | 2,225 (13%) | 5,773 (14%) | 76 (7.6%) |
| North West | 12,930 (17%) | 2,178 (14%) | 3,296 (19%) | 7,311 (17%) | 145 (15%) |
| Scotland | 3,068 (4.1%) | 606 (4.0%) | 573 (3.4%) | 1,846 (4.4%) | 43 (4.3%) |
| South East | 9,450 (13%) | 2,131 (14%) | 1,974 (12%) | 5,052 (12%) | 293 (29%) |
| South West | 3,919 (5.2%) | 726 (4.8%) | 795 (4.7%) | 2,361 (5.6%) | 37 (3.7%) |
| Wales | 3,623 (4.8%) | 713 (4.7%) | 893 (5.3%) | 1,991 (4.7%) | 26 (2.6%) |

<sup>1</sup>Statistics presented: median (IQR); n (%)

### Development of the 4C Deterioration model

Since we hypothesised that geographic heterogeneity may contribute to model performance, we analysed the dataset by UK National Health Service (NHS) region. We included eight regions in model development (sample sizes range 3,068 to 15,599; total n=66,764), with one region (London; n=8,252) held out for additional validation. Candidate predictors and their proportions of missingness, stratified by NHS region, are shown in Supplementary Figure 2. Proportions of missingness appeared similar across regions for each variable.

Following our backward elimination procedure, 11 predictors were retained in >50% of multiply imputed datasets in >50% of NHS regions in the development cohort. These were: age, sex, nosocomial infection, Glasgow coma scale, admission oxygen saturation, breathing room air or oxygen therapy, respiratory rate, urea, C-reactive protein, lymphocyte count and presence of radiographic chest infiltrates. Associations (including non-linearities) between these predictors and the outcome from the model trained on the full development cohort are shown visually in Figure 1. Full model coefficients are presented to enable independent model reconstruction in Supplementary Table 2.

**Figure 1:**
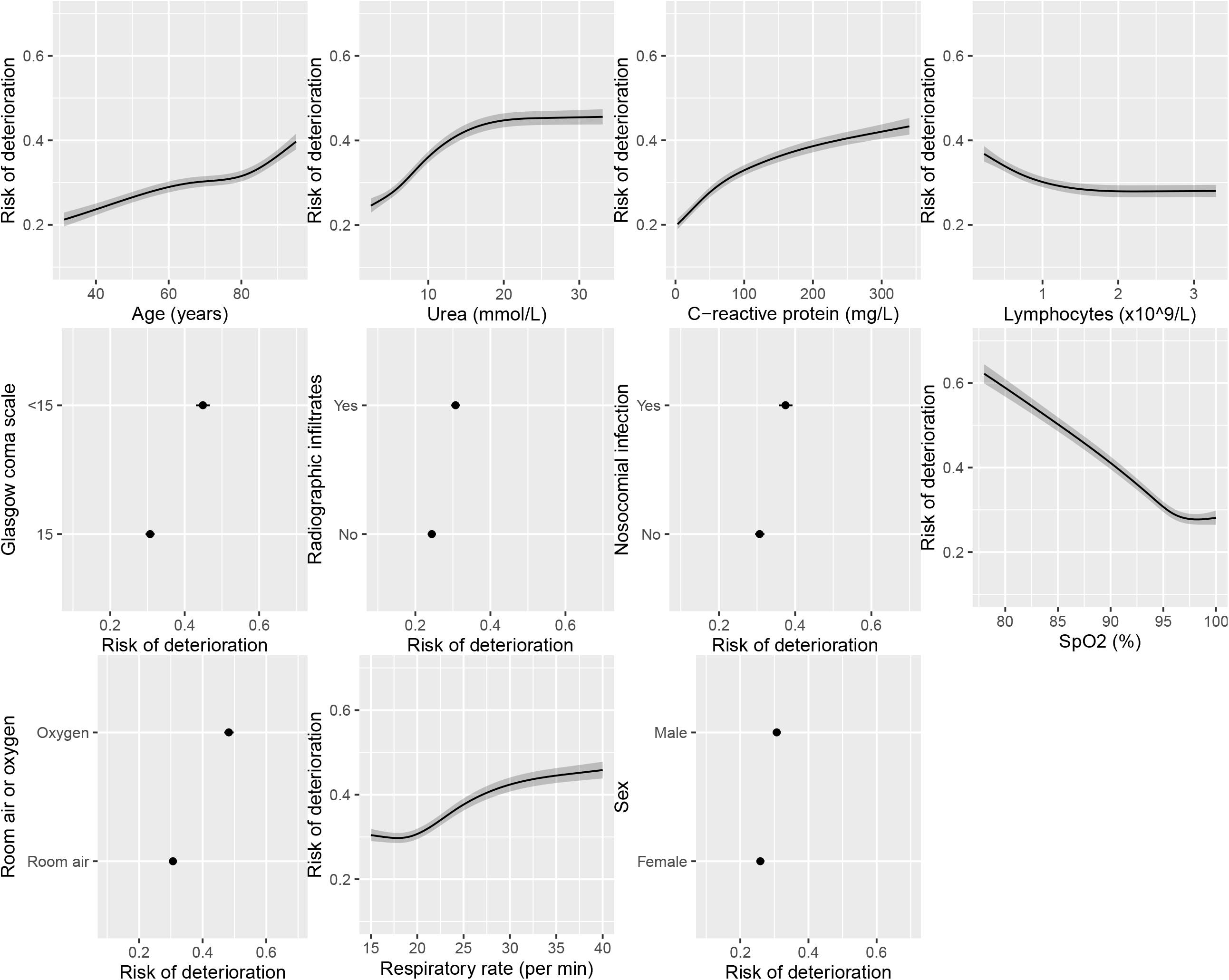
Multivariable associations between selected predictors and outcome in final model. Variable selection was done in each imputed dataset using backward elimination within each NHS region using AIC. Variables retained in >50% of multiply imputed datasets in >50% of NHS regions were selected. Continuous variables are modelled using restricted cubic splines. Final model parameters are pooled across multiply imputed datasets (total sample size for model development = 66764 participants). Black lines indicate point estimates; grey shaded regions indicate 95% confidence intervals.

**Table 2:**
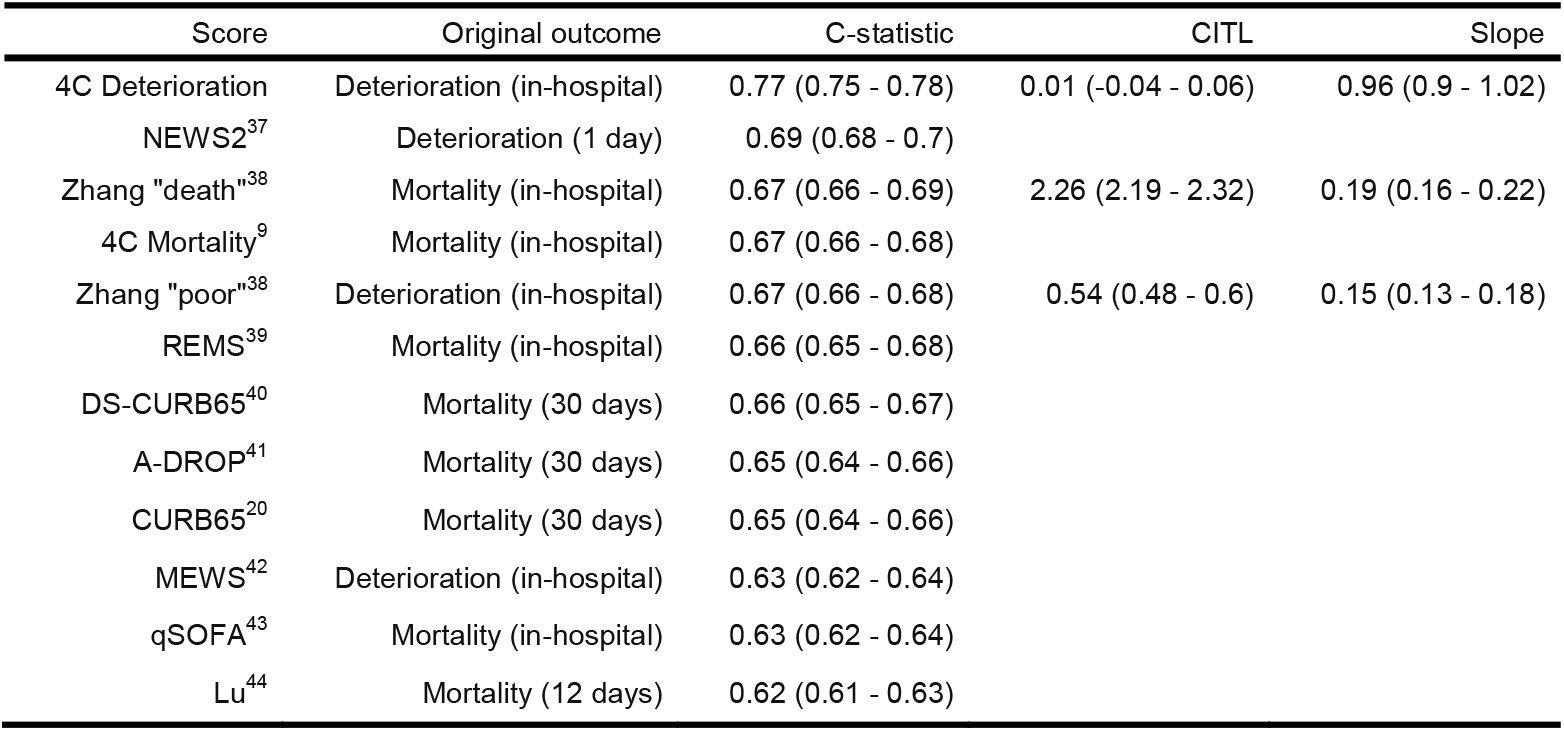
Validation performance in held-out London region. Models are shown for prediction of in-hospital clinical deterioration and are sorted by C-statistic (total sample size = 8252 participants). CITL = calibration-in-the-large. ‘Original outcome’ column indicates original intended outcome for each candidate model during development. CITL and slopes are not shown for points score models since they are not on probability scale.

### Internal-external cross-validation

In order to examine potential heterogeneity between NHS regions and evaluate generalisability, we conducted internal-external cross-validation^26^ of the prognostic model in the development cohort. In this process, we iteratively excluded one of the eight contributing NHS regions from the development set; the model was then trained using the 11 selected predictors in the remaining 7 development regions before being validated in the omitted region.

Forest plots showing model discrimination (C-statistic) and calibration metrics (slope and calibration-in-the-large (CITL) from internal-external cross-validation are shown in Figure 2. An ideal calibration slope is 1, while CITL should be 0 if the number of observed outcome events matches the number predicted. C-statistics were consistent across development NHS regions (point estimates 0.75-0.77; pooled random effects meta-analysis estimate 0.76; 95% confidence interval (95% CI) 0.75 to 0.77). Calibration slopes were also consistent across regions, with little evidence of heterogeneity (point estimates 0.95-1.06; pooled estimate 0.99; 95% CI 0.97 to 1.02). There was minor heterogeneity across NHS regions in CITL, likely reflecting some variation in baseline risk between regions (point estimates −0.20 to 0.13; pooled estimate −0.01; 95% CI −0.11 to 0.09). Overall risk was slightly underestimated in South East England (CITL 0.09; 95% CI 0.04 to 0.14) and Wales (CITL 0.13; 95% CI 0.06 to 0.21), and overestimated in Scotland (CITL −0.20; 95% CI −0.28 to −0.11) and South West England (CITL −0.19; 95% CI −0.26 to −0.11). Pooled calibration plots by NHS region are shown in Supplementary Figure 3a.

**Figure 2:**
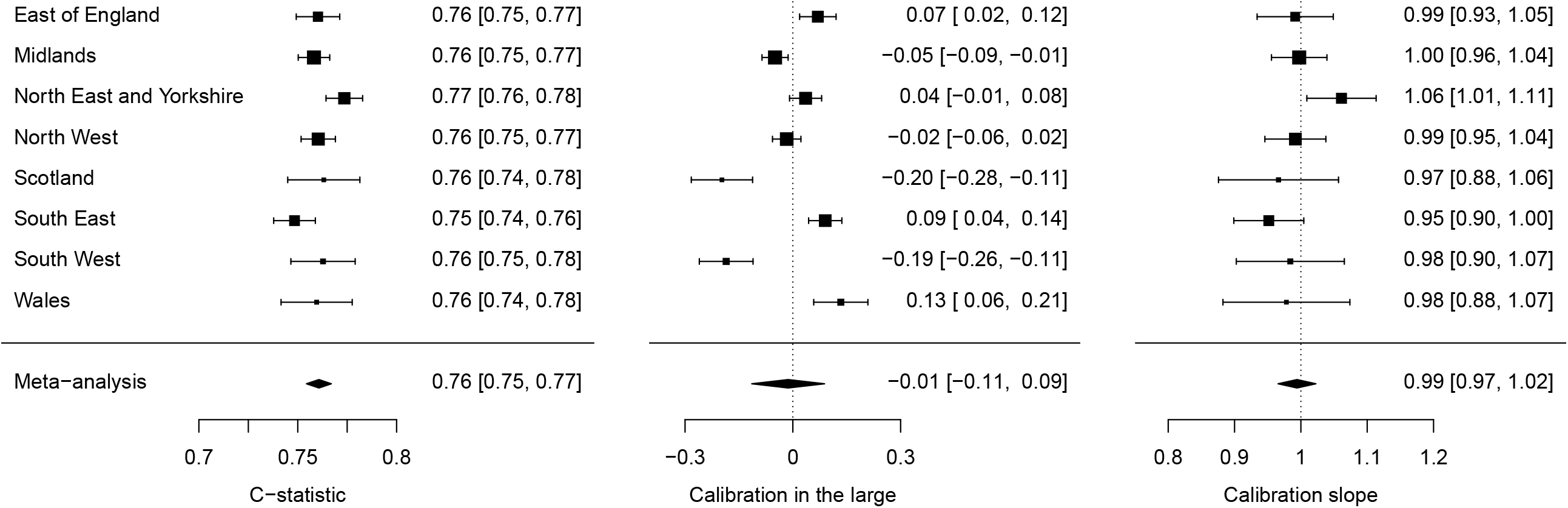
Internal-external cross validation of selected model by NHS region. Pooled estimates are calculated through random-effects meta-analysis (total sample size = 66,764 participants). Dashed lines indicate lines of perfect calibration in the large (0) and slope (1), respectively. Black squares indicate point estimates; bars indicate 95% confidence intervals; diamonds indicate pooled random-effects meta-analysis estimates.

In view of the minor variation in CITL between NHS regions, we also repeated internal-external cross validation with recalibration to each NHS region by re-estimation of the model intercept; calibration plots with recalibrated intercepts confirmed small improvements in model calibration (Supplementary Figure 3b).

Decision curve analyses in the validation sets in internal-external cross-validation, without recalibration of the new model, are shown in Supplementary Figure 4, with benchmarking to 11 existing candidate prognostic models for which the constituent variables were available in >60% of participants in our data. In decision curve analysis, net benefit allows assessment of clinical utility by quantifying the trade-off between correctly identifying true positives and incorrectly identifying false positives weighted according to the threshold probability^28^. The threshold probability represents the risk cut-off above which any given treatment or intervention might be considered, and reflects the underlying risk:benefit ratio for the intervention. The new model for clinical deterioration had higher net benefit than any of the existing models as well as ‘treat all’ or ‘treat none’ strategies, across a broad range of threshold probabilities, in all development NHS regions (without local recalibration).

### Validation in held-out NHS region

Next, we validated the final prognostic model, trained on the full development cohort, in the held-out NHS region (London; n=8,252). Discrimination and calibration metrics for the 4C Deterioration model were similar to the estimates from internal-external cross validation (Table 2), with C-statistic 0.77 (95% CI 0.75 to 0.78), CITL 0.01 (−0.04 to 0.06) and slope 0.96 (0.90 to 1.02). Discrimination was higher for the 4C Deterioration model than for the other existing candidates. A loess-smoothed calibration curve for the held-out London region is shown in Figure 3a.

We then conducted decision curve analysis in the held-out NHS region to further examine clinical utility for the 4C Deterioration model. Importantly, this demonstrated higher net benefit than all other candidates that we were able to recreate, as well as the ‘treat all’ and ‘treat none’ approaches, across a range of threshold probabilities (Figure 3b).

**Figure 3:**
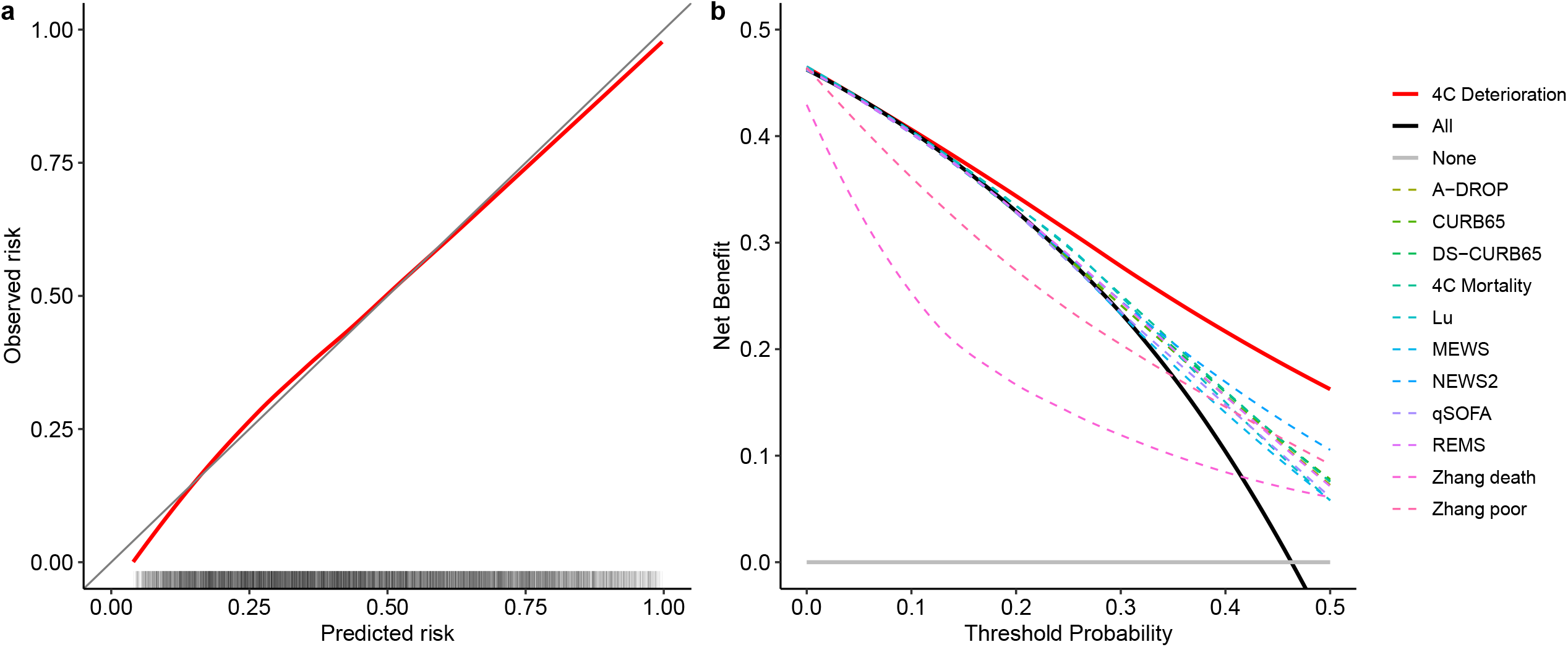
Calibration and decision curve analysis in held-out London region. Calibration (a) is shown using a loess-smoother across multiply imputed datasets. The rug plot indicates the distribution of predicted risk. Net benefit (b) is shown with loess smoothing for each candidate model compared to the ‘treat all’ and ‘treat none’ approaches. Points score models are recalibrated to the validation data, resulting in optimistic estimates of net benefit for these models. Total sample size = 8,252 participants).

We anticipate that clinicians may wish to evaluate risk of deterioration or death separately. Therefore, for illustration, we compared predictions from the 4C Deterioration model to our previously reported 4C Mortality Score^9^ in the London validation cohort, stratified by age (Figure 4a). In addition, 10 example participants selected at random from each decile of 4C Deterioration predictions in the London cohort are shown in Figure 4b, with their clinical characteristics summarised in Figure 4c. Overall, deterioration predictions appeared appropriately higher than those for mortality, but these differences were exaggerated among younger age groups.

**Figure 4:**
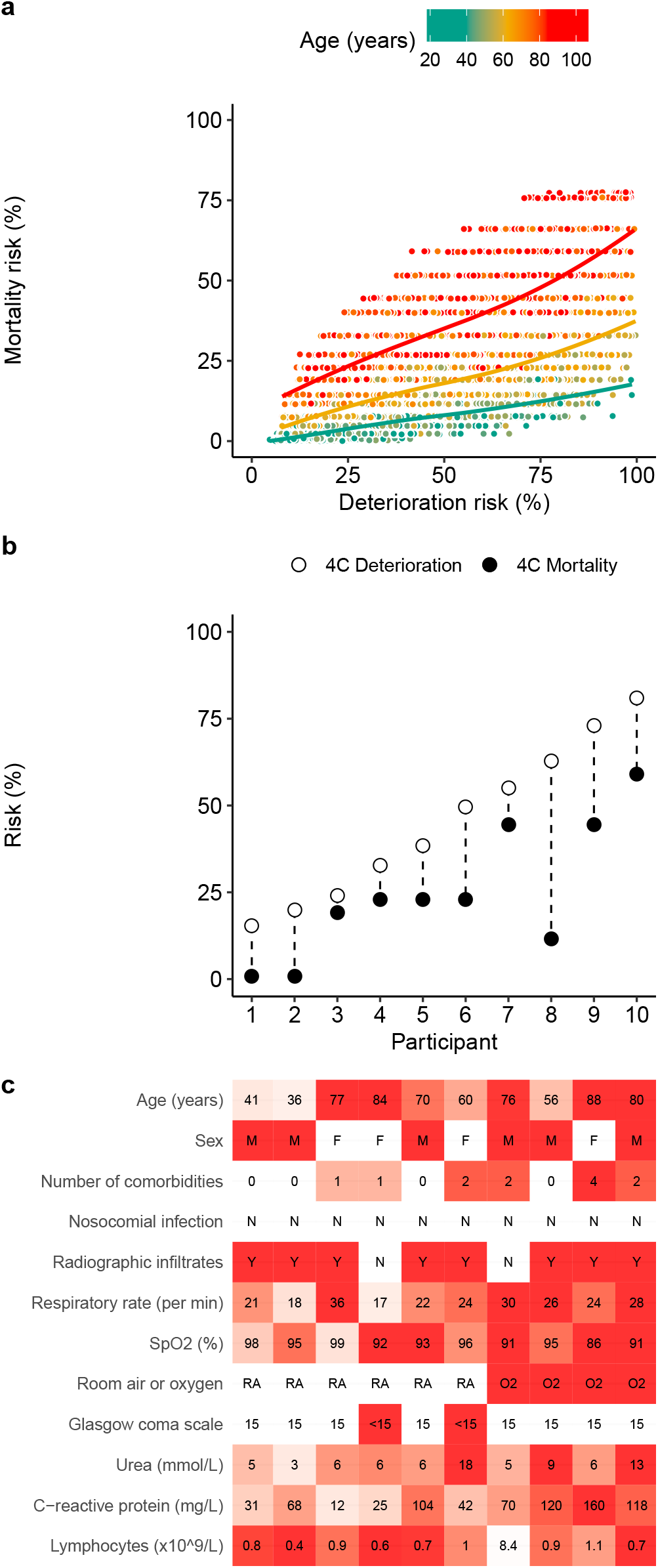
4C Deterioration vs Mortality predictions for (a) London validation cohort (n = 8252) and (b-c) randomly sampled example patients. 4C Mortality probabilities are calculated from points scores, based on observed mortality risk for each score in the original validation data. In panel (a) smoothed plot reflects loess fit, stratified by age (green = under 50 years; yellow = 50 to 69; red = 70 or older). In panel (b), example patients are randomly sampled from the validation cohort, stratified by deciles of 4C Deterioration model predictions. Characteristics of each example participant are shown in (c), with red indicating characteristics associated with higher risk predictions. In (c), M = male; F = female; O2 = receiving oxygen therapy; RA = room air; Y = yes; N = no.

### Sensitivity analyses

Recalculation of model validation metrics in complete case data from the held-out London region showed similar results to the primary analyses (Supplementary Table 3). Exclusion of deterioration events on the day of admission in the London validation cohort resulted in slightly lower C-statistics for most models; discrimination remained higher for the 4C Deterioration model, compared to other candidates (Supplementary Table 4). Validation metrics in the London cohort appeared similar to the primary analysis when excluding participants who had ongoing hospital care at the end of follow-up (Supplementary Table 5), when restricted to community-acquired infections (Supplementary Table 6a), and when community-acquired infections with symptom onset after admission were excluded (Supplementary Table 7). Among nosocomial infections, the C-statistic appeared slightly lower for the 4C Deterioration model than in the primary analysis (0.72; 95% CI 0.67 to 0.77), though discrimination remained higher than other candidate models, and CITL was 0.39 (95% CI 0.2 to 0.59), suggesting some underestimation of risk (Supplementary Table 6b). Repeating the analyses using an alternative multiple imputation approach and with shorter and longer temporal definitions of nosocomial infection led to similar results to the primary analysis (Supplementary Figures 5-8; Supplementary Table 8).

## Discussion

In this study, we developed and validated a prognostic model for clinical deterioration in a population of 75,016 consecutive adults hospitalised with COVID-19 and recruited to the ISARIC4C study across 260 hospitals in England, Scotland and Wales. The final model integrates 11 predictors that are routinely measured in clinical practice. The model was initially validated through internal-external cross-validation, demonstrating consistent discrimination, calibration and net benefit across NHS regions. The final model was then further validated in the held-out London region and demonstrated consistent performance. Importantly, the 4C Deterioration model achieved higher net benefit than other candidate risk stratification tools across a broad range of risk thresholds - in all NHS regions. Thus, the 4C Deterioration model demonstrates strong potential for clinical utility and generalisability.

Our 4C Deterioration model can be implemented programmatically alongside our previously reported 4C Mortality Score^9^. The comparison of 4C Deterioration *vs*. 4C Mortality predictions among the London validation cohort showed higher predictions for deterioration overall; these differences were amplified among younger age groups. This suggests that younger people who deteriorated were more likely to have escalation of treatment through HDU/ICU admission or ventilatory support, while older people who deteriorated were more likely to die. These observations are likely to be mediated, in part, by differential treatment escalation decisions by age. Moreover, our comparison of the models for 10 randomly selected patients across the distribution of outcome risks from the held-out validation cohort illustrates examples of cases with relatively low risk of death, but moderate to high risk of deterioration. These discordances underline the need for independent prognostic models for deterioration and mortality outcomes, thus empowering clinicians to predict their desired outcome as required to inform clinical management decisions.

Our study has a number of strengths. Previous studies seeking to develop prognostic models for people with COVID-19 have been evaluated as being at high risk of bias due to suboptimal development methodology, and are often limited to single hospital sites^6^, thus impeding generalisability during external validation^8^. In the current study, we adhered to TRIPOD standards^11^ and retained continuous variables without arbitrary categorisation, while accounting for linearities, to avoid loss of information^32^. Moreover, we used the largest dataset to date, to our knowledge, to develop and validate the 4C Deterioration model, including data from hospitals across nine NHS regions in England, Scotland and Wales. We exploited this wide geographic coverage to explore between-region heterogeneity in model performance using the recommended approach of internal-external cross-validation^33^. While discrimination, calibration slopes and net benefit were largely very consistent, we noted minor variation in CITL, suggesting some variation in baseline risk between regions. Our approach of recalibrating the model intercept to each NHS region demonstrated the potential to address such heterogeneity and could be used to update the model if risk is found to vary temporally (as novel therapies are implemented) and among different populations. However, it is notable that net benefit, which accounts for model discrimination and calibration in quantifying clinical utility, appeared higher for the 4C Deterioration model than all other candidates even without recalibration, across all NHS regions and in the held-out validation dataset. This was the case even when comparing to points-based models, which may achieve overly optimistic performance in during decision curve analysis since they were recalibrated to the validation data set. We also used a robust approach to missing data with multiple imputation, as widely recommended in prediction model studies^34^, and performed a sensitivity analysis using an alternative approach, with similar findings.

Ongoing prospective external validation of the 4C Deterioration model will be required over time to consider the need for temporal recalibration^35^, and to include diverse international settings outside of the ISARIC4C study. Another limitation is that we only included predictors that were routinely measured as part of clinical care during the study period, and specified that they had to be available among >60% of the population for inclusion in the analysis. Thus, we were unable to assess candidate models that include predictors such as lactate dehydrogenase or D-dimer, since these variables were only available in a small minority of participants. Future studies could consider standardised capture of laboratory measurements considered to have prognostic value to enable inclusion of these variables in model development and validation at scale. Moreover, we note that novel molecular biomarkers currently under investigation may also offer prognostic value^36^. Blood transcript, protein and metabolite measurements will be available from a subset of the ISARIC4C participants and could be integrated into risk-stratification tools in future studies.

In summary, we present a prognostic model for clinical deterioration among hospitalised adults with community or hospital acquired COVID-19, validated in nine NHS regions in England, Scotland and Wales. The model uses readily available clinical predictors and will be made freely available online alongside our previously reported mortality risk score (https://isaric4c.net/outputs/4c_score/)^9^ at the point of peer-reviewed publication, to inform clinical decision-making and patient stratification for therapeutic interventions. The underlying model coefficients are presented and code will be published to enable independent external validation in new datasets.

## Supporting information

TRIPOD checklist

## Data Availability

We welcome applications for data and material access via our Independent Data and Material Access Committee (https://isaric4c.net).

## Footnotes

### ISARIC4C Investigators

<u>Consortium Lead Investigator:</u> J Kenneth Baillie, Chief Investigator: Malcolm G Semple, Co-Lead Investigator: Peter JM Openshaw. <u>ISARIC Clinical Coordinator:</u> Gail Carson. <u>Co-Investigators:</u> Beatrice Alex, Benjamin Bach, Wendy S Barclay, Debby Bogaert, Meera Chand, Graham S Cooke, Annemarie B Docherty, Jake Dunning, Ana da Silva Filipe, Tom Fletcher, Christopher A Green, Ewen M Harrison, Julian A Hiscox, Antonia Ying Wai Ho, Peter W Horby, Samreen Ijaz, Saye Khoo, Paul Klenerman, Andrew Law, Wei Shen Lim, Alexander J Mentzer, Laura Merson, Alison M Meynert, Mahdad Noursadeghi, Shona C Moore, Massimo Palmarini, William A Paxton, Georgios Pollakis, Nicholas Price, Andrew Rambaut, David L Robertson, Clark D Russell, Vanessa Sancho-Shimizu, Janet T Scott, Thushan de Silva, Louise Sigfrid, Tom Solomon, Shiranee Sriskandan, David Stuart, Charlotte Summers, Richard S Tedder, Emma C Thomson, AA Roger Thompson, Ryan S Thwaites, Lance CW Turtle, Maria Zambon. Project Managers: Hayley Hardwick, Chloe Donohue, Ruth Lyons, Fiona Griffiths, Wilna Oosthuyzen. <u>Data Analysts:</u> Lisa Norman, Riinu Pius, Tom M Drake, Cameron J Fairfield, Stephen Knight, Kenneth A Mclean, Derek Murphy, Catherine A Shaw. <u>Data and Information System Managers:</u> Jo Dalton, James Lee, Daniel Plotkin, Michelle Girvan, Egle Saviciute, Stephanie Roberts, Janet Harrison, Laura Marsh, Marie Connor, Sophie Halpin, Clare Jackson, Carrol Gamble. <u>Data integration and presentation:</u> Gary Leeming, Andrew Law, Murray Wham, Sara Clohisey, Ross Hendry, James Scott-Brown. <u>Material Management:</u> William Greenhalf, Victoria Shaw, Sarah McDonald. <u>Patient engagement:</u> Seán Keating. <u>Outbreak Laboratory Staff and Volunteers:</u> Katie A. Ahmed, Jane A Armstrong, Milton Ashworth, Innocent G Asiimwe, Siddharth Bakshi, Samantha L Barlow, Laura Booth, Benjamin Brennan, Katie Bullock, Benjamin WA Catterall, Jordan J Clark, Emily A Clarke, Sarah Cole, Louise Cooper, Helen Cox, Christopher Davis, Oslem Dincarslan, Chris Dunn, Philip Dyer, Angela Elliott, Anthony Evans, Lorna Finch, Lewis WS Fisher, Terry Foster, Isabel Garcia-Dorival, Willliam Greenhalf, Philip Gunning, Catherine Hartley, Antonia Ho, Rebecca L Jensen, Christopher B Jones, Trevor R Jones, Shadia Khandaker, Katharine King, Robyn T. Kiy, Chrysa Koukorava, Annette Lake, Suzannah Lant, Diane Latawiec, L Lavelle-Langham, Daniella Lefteri, Lauren Lett, Lucia A Livoti, Maria Mancini, Sarah McDonald, Laurence McEvoy, John McLauchlan, Soeren Metelmann, Nahida S Miah, Joanna Middleton, Joyce Mitchell, Shona C Moore, Ellen G Murphy, Rebekah Penrice-Randal, Jack Pilgrim, Tessa Prince, Will Reynolds, P. Matthew Ridley, Debby Sales, Victoria E Shaw, Rebecca K Shears, Benjamin Small, Krishanthi S Subramaniam, Agnieska Szemiel, Aislynn Taggart, Jolanta Tanianis-Hughes, Jordan Thomas, Erwan Trochu, Libby van Tonder, Eve Wilcock, J. Eunice Zhang. <u>Local Principal Investigators:</u> Kayode Adeniji, Daniel Agranoff, Ken Agwuh, Dhiraj Ail, Ana Alegria, Brian Angus, Abdul Ashish, Dougal Atkinson, Shahedal Bari, Gavin Barlow, Stella Barnass, Nicholas Barrett, Christopher Bassford, David Baxter, Michael Beadsworth, Jolanta Bernatoniene, John Berridge, Nicola Best, Pieter Bothma, David Brealey, Robin Brittain-Long, Naomi Bulteel, Tom Burden, Andrew Burtenshaw, Vikki Caruth, David Chadwick, Duncan Chambler, Nigel Chee, Jenny Child, Srikanth Chukkambotla, Tom Clark, Paul Collini, Catherine Cosgrove, Jason Cupitt, Maria-Teresa Cutino-Moguel, Paul Dark, Chris Dawson, Samir Dervisevic, Phil Donnison, Sam Douthwaite, Ingrid DuRand, Ahilanadan Dushianthan, Tristan Dyer, Cariad Evans, Chi Eziefula, Chrisopher Fegan, Adam Finn, Duncan Fullerton, Sanjeev Garg, Sanjeev Garg, Atul Garg, Effrossyni Gkrania-Klotsas, Jo Godden, Arthur Goldsmith, Clive Graham, Elaine Hardy, Stuart Hartshorn, Daniel Harvey, Peter Havalda, Daniel B Hawcutt, Maria Hobrok, Luke Hodgson, Anil Hormis, Michael Jacobs, Susan Jain, Paul Jennings, Agilan Kaliappan, Vidya Kasipandian, Stephen Kegg, Michael Kelsey, Jason Kendall, Caroline Kerrison, Ian Kerslake, Oliver Koch, Gouri Koduri, George Koshy, Shondipon Laha, Steven Laird, Susan Larkin, Tamas Leiner, Patrick Lillie, James Limb, Vanessa Linnett, Jeff Little, Michael MacMahon, Emily MacNaughton, Ravish Mankregod, Huw Masson, Elijah Matovu, Katherine McCullough, Ruth McEwen, Manjula Meda, Gary Mills, Jane Minton, Mariyam Mirfenderesky, Kavya Mohandas, Quen Mok, James Moon, Elinoor Moore, Patrick Morgan, Craig Morris, Katherine Mortimore, Samuel Moses, Mbiye Mpenge, Rohinton Mulla, Michael Murphy, Megan Nagel, Thapas Nagarajan, Mark Nelson, Igor Otahal, Mark Pais, Selva Panchatsharam, Hassan Paraiso, Brij Patel, Natalie Pattison, Justin Pepperell, Mark Peters, Mandeep Phull, Stefania Pintus, Jagtur Singh Pooni, Frank Post, David Price, Rachel Prout, Nikolas Rae, Henrik Reschreiter, Tim Reynolds, Neil Richardson, Mark Roberts, Devender Roberts, Alistair Rose, Guy Rousseau, Brendan Ryan, Taranprit Saluja, Aarti Shah, Prad Shanmuga, Anil Sharma, Anna Shawcross, Jeremy Sizer, Manu Shankar-Hari, Richard Smith, Catherine Snelson, Nick Spittle, Nikki Staines, Tom Stambach, Richard Stewart, Pradeep Subudhi, Tamas Szakmany, Kate Tatham, Jo Thomas, Chris Thompson, Robert Thompson, Ascanio Tridente, Darell Tupper-Carey, Mary Twagira, Andrew Ustianowski, Nick Vallotton, Lisa Vincent-Smith, Shico Visuvanathan, Alan Vuylsteke, Sam Waddy, Rachel Wake, Andrew Walden, Ingeborg Welters, Tony Whitehouse, Paul Whittaker, Ashley Whittington, Meme Wijesinghe, Martin Williams, Lawrence Wilson, Sarah Wilson, Stephen Winchester, Martin Wiselka, Adam Wolverson, Daniel G Wooton, Andrew Workman, Bryan Yates, and Peter Young.

## Acknowledgements

This work uses data provided by patients and collected by the NHS as part of their care and support #DataSavesLives. We are extremely grateful to the 2,648 frontline NHS clinical and research staff and volunteer medical students, who collected this data in challenging circumstances; and the generosity of the participants and their families for their individual contributions in these difficult times. We also acknowledge the support of Jeremy J Farrar, Nahoko Shindo, Devika Dixit, Nipunie Rajapakse, Lyndsey Castle, Martha Buckley, Debbie Malden, Katherine Newell, Kwame O’Neill, Emmanuelle Denis, Claire Petersen, Scott Mullaney, Sue MacFarlane, Nicole Maziere, Julien Martinez, Oslem Dincarslan, and Annette Lake.

## Author contributions

Conceptualisation: RKG, EMH, AH, ABD, PJMO, JKB, MGS, MN;

Data curation: RP, SH, KAH, CJ, CAG, KAM, LM, LN, CDR, CAS, MGS, ABD, EMH;

Formal analysis: RKG, EMH, MN;

Funding acquisition: PWH, JSN-V-T, TS, PJMO, JKB, MGS, ABD;

Investigation: RKG, EMH, AH, ABD, PJMO, JKB, MGS, MN;

Methodology: RKG, EMH, AH, ABD, MVS, IA, ML, MQ, PJMO, JKB, MGS, MN;

Project administration: SH, HEH, CJ, LM, LN, CDR, CAS, MGS; Resources: CG, PWH, LM, TS, MGS, EMH;

Software: N/A;

Supervision: RKG, EMH, AH, ABD, IA, ML, PJMO, JKB, MGS, MN;

Visualisation: RKG, EMH, JS-B, MN; Writing-original draft: RKG, MN;

Writing-review and editing: RKG, EMH, AH, ABD, MVS, IA, ML, MQ, SRK, RP, IB, GC, TMD, CAG, JD, CJF, LM, JSN-V-T, PLO, MGP, CDR, JS-B, AS, CS, OVS, LCWT, PJMO, JKB, MGS, MN.

## Code availability statement

The final prognostic model developed in this study will be made freely available at the point of peer-reviewed publication, to enable implementation in clinical practice and independent external validation in new datasets. The code underlying the prediction tool will also be made available.

## References

1. World Health Organization. Coronavirus disease 2019 (COVID-19) Weekly Epidemiological Update. https://www.who.int/emergencies/diseases/novel-coronavirus-2019/situation-reports (2020).

2. Furlow, B. COVACTA trial raises questions about tocilizumab’s benefit in COVID-19. Lancet Rheumatol. (2020) doi:10.1016/S2665-9913(20)30313-1.

3. Beigel, J. H. et al. Remdesivir for the Treatment of Covid-19 — Preliminary Report. N. Engl. J. Med. NEJMoa2007764 (2020) doi:10.1056/NEJMoa2007764.

4. Sterne, J. A. C. et al. Association Between Administration of Systemic Corticosteroids and Mortality Among Critically Ill Patients With COVID-19. JAMA (2020) doi:10.1001/jama.2020.17023.

5. Dexamethasone in Hospitalized Patients with Covid-19 — Preliminary Report. N. Engl. J. Med. NEJMoa2021436 (2020) doi:10.1056/NEJMoa2021436.

6. Wynants, L. et al. Prediction models for diagnosis and prognosis of covid-19: systematic review and critical appraisal. BMJ 369, (2020).

7. Wolff, R. F. et al. PROBAST: A Tool to Assess the Risk of Bias and Applicability of Prediction Model Studies. Ann. Intern. Med. 170, 51 (2019).

8. Gupta, R. K. et al. Systematic evaluation and external validation of 22 prognostic models among hospitalised adults with COVID-19: An observational cohort study. Eur. Respir. J. (2020) doi:10.1183/13993003.03498-2020.

9. Knight, S. R. et al. Risk stratification of patients admitted to hospital with covid-19 using the ISARIC WHO Clinical Characterisation Protocol: development and validation of the 4C Mortality Score. BMJ m3339 (2020) doi:10.1136/bmj.m3339.

10. Docherty, A. B. et al. Features of 20□133 UK patients in hospital with covid-19 using the ISARIC WHO Clinical Characterisation Protocol: prospective observational cohort study. BMJ 369, m1985.(2020).

11. Collins, G. S., Reitsma, J. B., Altman, D. G. & Moons, K. G. M. Transparent reporting of a multivariable prediction model for individual prognosis or diagnosis (TRIPOD): the TRIPOD statement. BMJ 350, (2015).

12. COVID-19 Clinical Research Resources-ISARIC. https://isaric.tghn.org/covid-19-clinical-research-resources/.

13. NHS Regional Teams. “https://www.england.nhs.uk/about/regional-area-teams/”.

14. WHO Working Group on the Clinical Characterisation and Management of COVID-19 infection, J. C. et al. A minimal common outcome measure set for COVID-19 clinical research. Lancet. Infect. Dis. 0, (2020).

15. European Centre for Disease Prevention and Control. Surveillance definitions for COVID-19. “https://www.ecdc.europa.eu/en/covid-19/surveillance/surveillance-definitions”.

16. Charlson, M. E., Pompei, P., Ales, K. L. & MacKenzie, C. R. A new method of classifying prognostic comorbidity in longitudinal studies: development and validation. J. Chronic Dis. 40, 373–83 (1987).

17. Frank E Harrell. Biostatistical Modeling. https://biostat.mc.vanderbilt.edu/wiki/pub/Main/BioMod/notes.pdf (2004).

18. Seymour, C. W. et al. Assessment of Clinical Criteria for Sepsis. JAMA 315, 762 (2016).

19. Royal College of Physicians. National Early Warning Score (NEWS) 2 | RCP London. “https://www.rcplondon.ac.uk/projects/outputs/national-early-warning-score-news-2”.

20. Lim, W. S. et al. Defining community acquired pneumonia severity on presentation to hospital: an international derivation and validation study. Thorax 58, 377–382 (2003).

21. Riley, R. D. et al. Minimum sample size for developing a multivariable prediction model: PART II-binary and time-to-event outcomes. Stat. Med. 38, 1276–1296 (2019).

22. Riley, R. D. et al. Calculating the sample size required for developing a clinical prediction model. BMJ 368, m441.(2020).

23. White, I. R., Royston, P. & Wood, A. M. Multiple imputation using chained equations: Issues and guidance for practice. Stat. Med. 30, 377–399 (2011).

24. Buuren, S. van & Groothuis-Oudshoorn, K. mice: Multivariate Imputation by Chained Equations in R. J. Stat. Softw. 45(3), 1–67 (2011).

25. Rubin, D. B. Multiple imputation for nonresponse in surveys. (Wiley-Interscience, 2004).

26. Debray, T. P. A. et al. Individual Participant Data (IPD) Meta-analyses of Diagnostic and Prognostic Modeling Studies: Guidance on Their Use. PLOS Med. 12, e1001886.(2015).

27. Steyerberg, E. W. & Vergouwe, Y. Towards better clinical prediction models: seven steps for development and an ABCD for validation. Eur. Heart J. 35, 1925–31 (2014).

28. Vickers, A. J., van Calster, B. & Steyerberg, E. W. A simple, step-by-step guide to interpreting decision curve analysis. Diagnostic Progn. Res. 3, 18 (2019).

29. Wickham, H. et al. Welcome to the Tidyverse. J. Open Source Softw. 4, 1686 (2019).

30. Harrell Jr, F. E. rms: Regression Modeling Strategies. (2019).

31. Brown, M. rmda: Risk Model Decision Analysis. (2018).

32. Royston, P., Altman, D. G. & Sauerbrei, W. Dichotomizing continuous predictors in multiple regression: a bad idea. Stat. Med. 25, 127–41 (2006).

33. Steyerberg, E. W. & Harrell, F. E. Prediction models need appropriate internal, internal– external, and external validation. J. Clin. Epidemiol. 69, 245–247 (2016).

34. Riley, R. D., Windt, D.van der Croft, P. & Moons, K. G. M. Prognosis research in healthcare□: concepts, methods, and impact.

35. Booth, S., Riley, R. D., Ensor, J., Lambert, P. C. & Rutherford, M. J. Temporal recalibration for improving prognostic model development and risk predictions in settings where survival is improving over time. Int. J. Epidemiol. (2020) doi:10.1093/ije/dyaa030.

36. Del Valle, D. M. et al. An inflammatory cytokine signature predicts COVID-19 severity and survival. Nat. Med. (2020) doi:10.1038/s41591-020-1051-9.

37. Smith, G. B., Prytherch, D. R., Meredith, P., Schmidt, P. E. & Featherstone, P. I. The ability of the National Early Warning Score (NEWS) to discriminate patients at risk of early cardiac arrest, unanticipated intensive care unit admission, and death. Resuscitation 84, 465–470 (2013).

38. Zhang, H. et al. Risk prediction for poor outcome and death in hospital in-patients with COVID-19: derivation in Wuhan, China and external validation in London, UK. medRxiv 2020.04.28.20082222 (2020) doi:10.1101/2020.04.28.20082222.

39. Olsson, T., Terent, A. & Lind, L. Rapid Emergency Medicine score: a new prognostic tool for in-hospital mortality in nonsurgical emergency department patients. J. Intern. Med. 255, 579–587 (2004).

40. Dwyer, R., Hedlund, J., Henriques-Normark, B. & Kalin, M. Improvement of CRB-65 as a prognostic tool in adult patients with community-acquired pneumonia. BMJ Open Respir. Res. 1, e000038.(2014).

41. Miyashita, N., Matsushima, T., Oka, M. & Japanese Respiratory Society. The JRS guidelines for the management of community-acquired pneumonia in adults: an update and new recommendations. Intern. Med. 45, 419–28 (2006).

42. Subbe, C. P., Kruger, M., Rutherford, P. & Gemmel, L. Validation of a modified Early Warning Score in medical admissions. QJM 94, 521–526 (2001).

43. Seymour, C. W. et al. Assessment of Clinical Criteria for Sepsis. JAMA 315, 762 (2016).

44. Lu, J. et al. ACP risk grade: a simple mortality index for patients with confirmed or suspected severe acute respiratory syndrome coronavirus 2 disease (COVID-19) during the early stage of outbreak in Wuhan, China. medRxiv 2020.02.20.20025510 (2020) doi:10.1101/2020.02.20.20025510.

