## Supplementary Information for "Development and validation of the 4C Deterioration model for adults hospitalised with COVID-19"

Supplementary Material

Supplementary Figure 1: Days to deterioration following admission, stratified by first chronological type of deterioration.

Outcome events are shown as stacked histogram. Total sample size = 75,016 participants with 30,939 deterioration events with known time point shown in plot. Median days to deterioration was 4 (interquartile range 1-9).

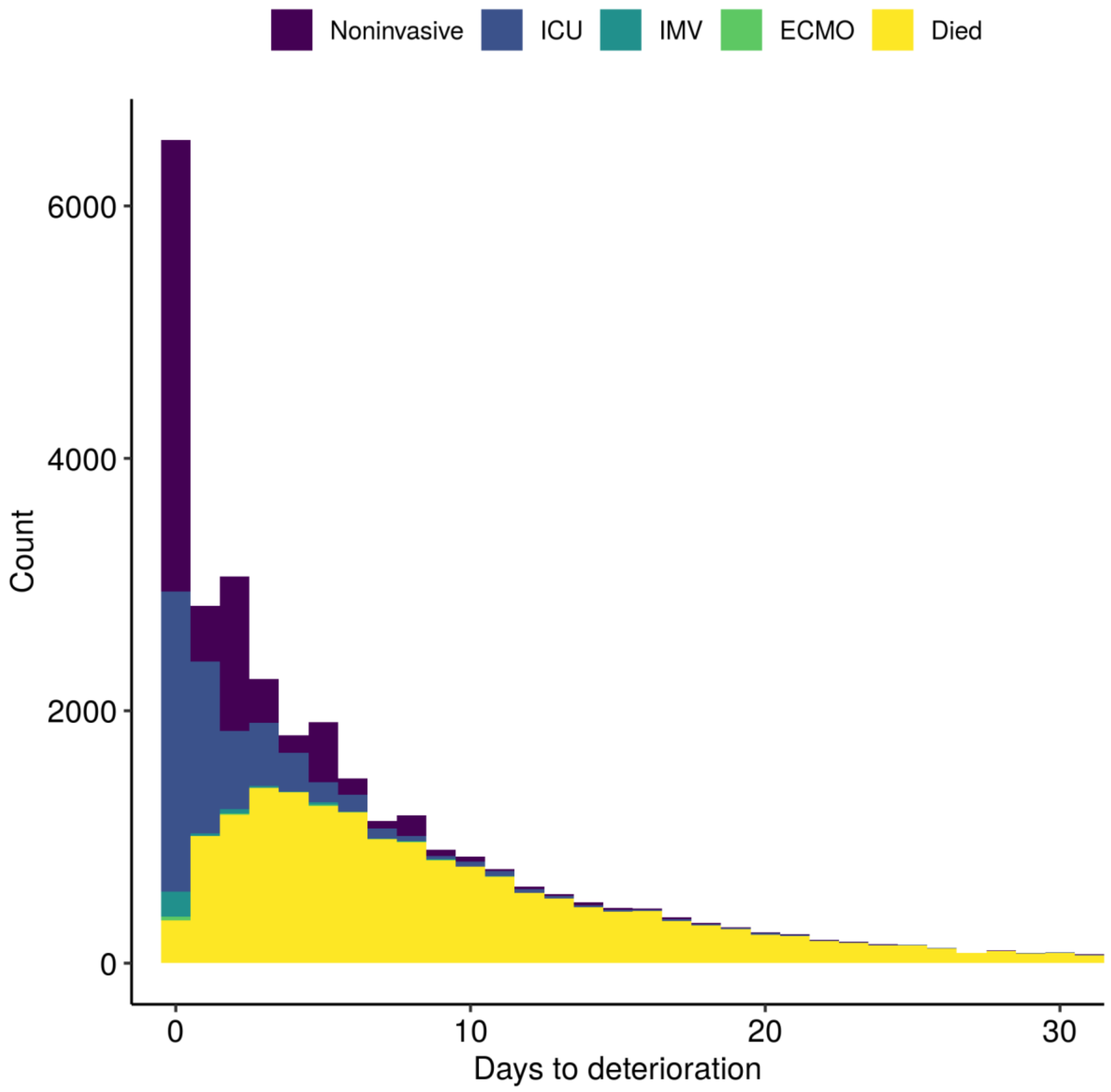

Supplementary Figure 2: Heatmap showing missingness (%) of candidate predictors and outcome, stratified by NHS region (n = 75,016 participants).

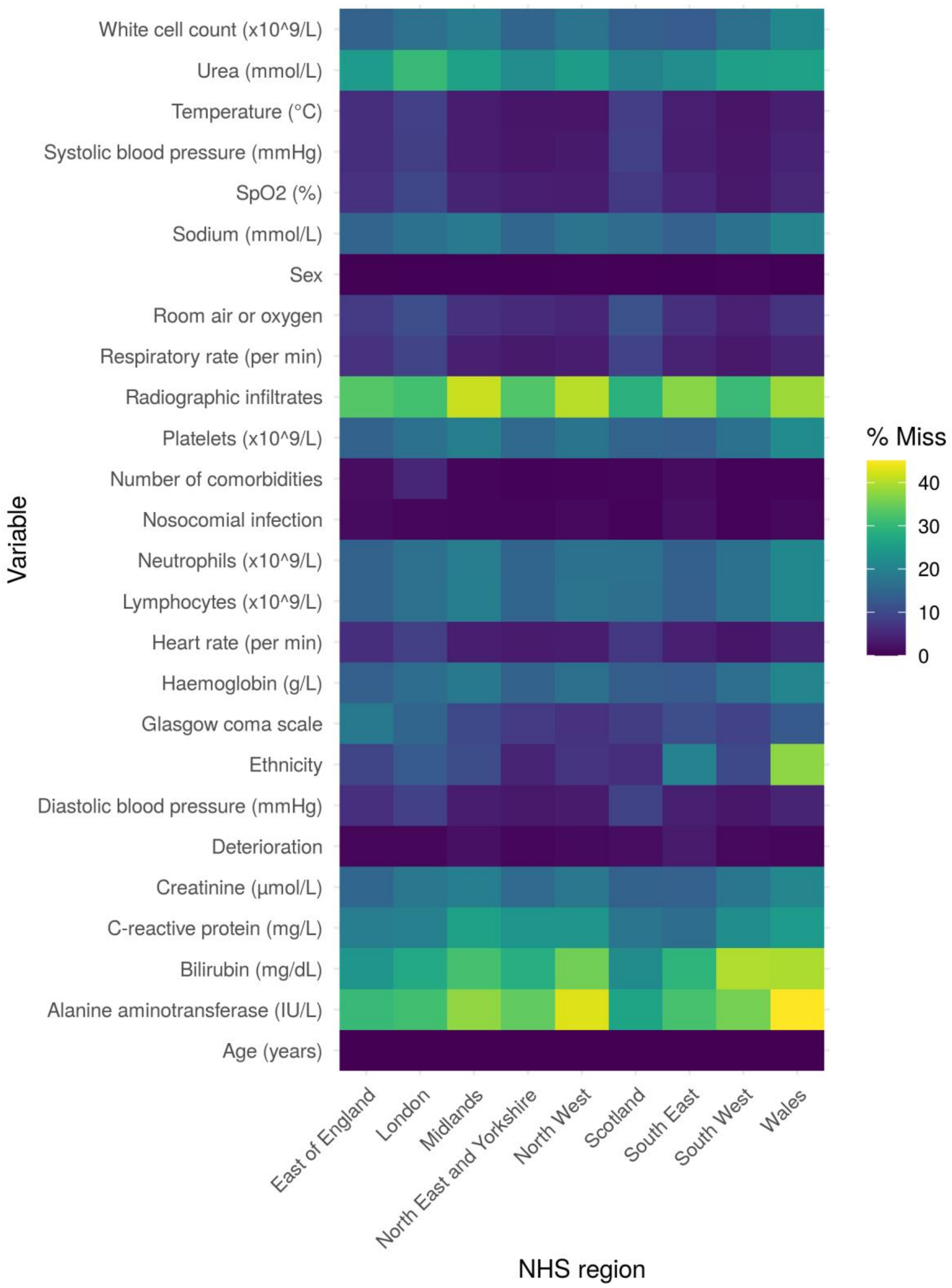

Supplementary Figure 3: Pooled calibration across multiple imputed datasets during IECV (a) before and (b) after recalibration to local regions.

Calibration is shown using a loess-smoother across multiply imputed datasets. Rug plots indicate the distributions of predicted risk. Total sample size = 66,764 participants.

(a)

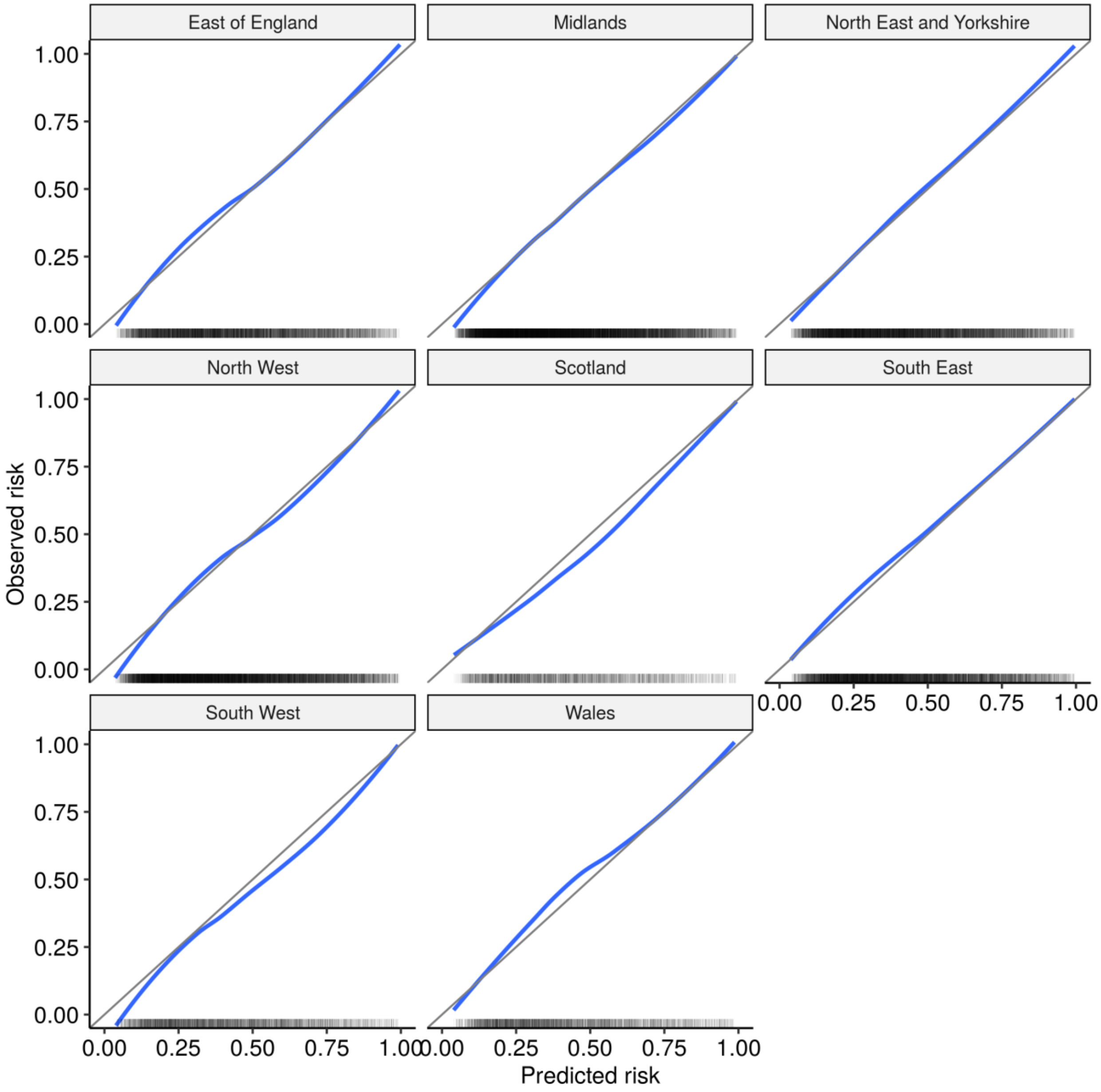

(b)

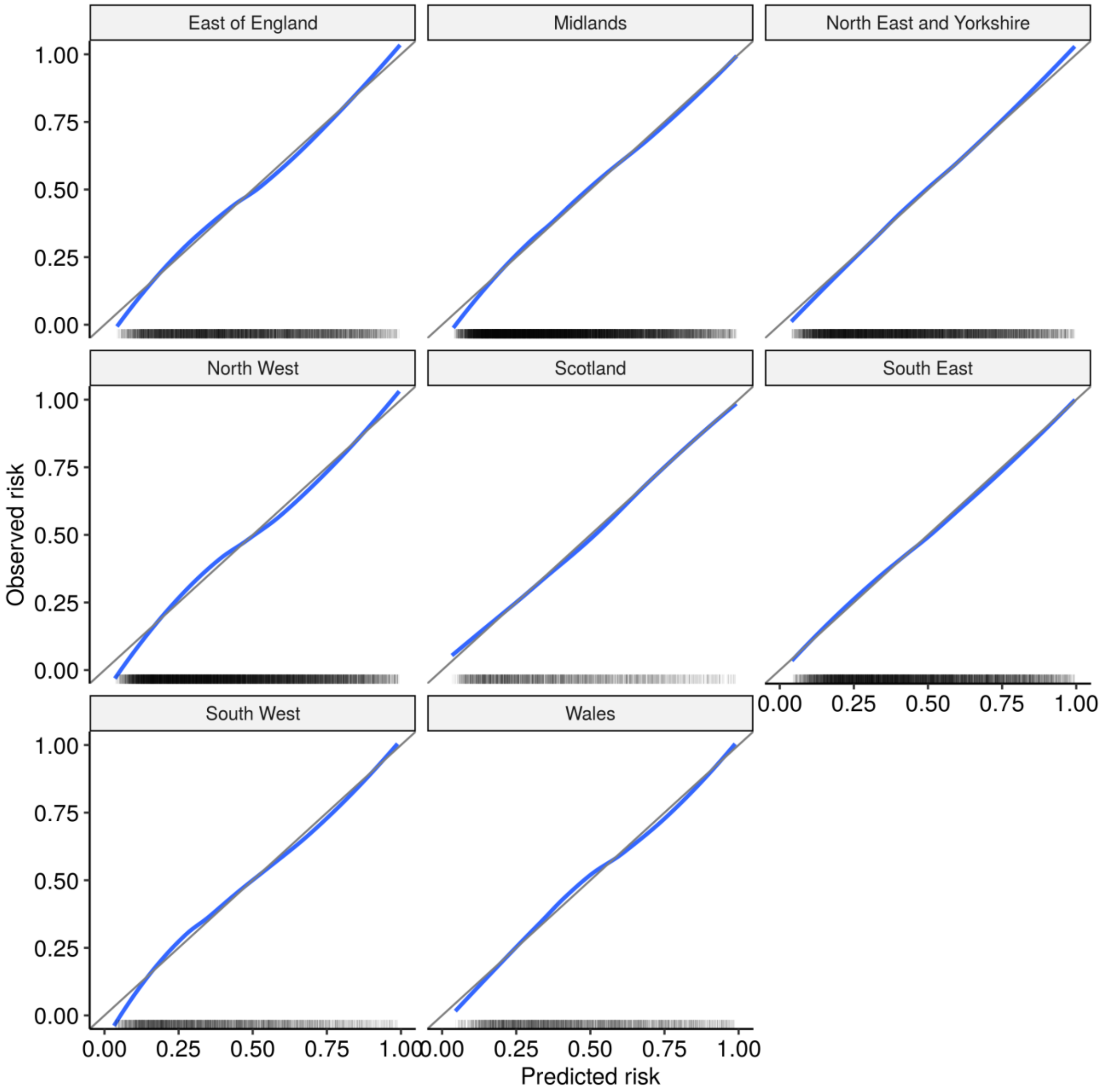
East of England
Midlands
North East and Yorkshire
North West
Scotland
South East
South West
Wales
Observed risk
Predicted risk
0.00
0.25
0.50
0.75
1.00

Supplementary Figure 4: Decision curve analysis during internal-external cross validation.

Shown for the 4C Deterioration model without recalibration to local region. Net benefit is shown for each candidate model compared to the ‘treat all’ and ‘treat none’ approaches. Points score models are recalibrated to the validation data, resulting in optimistic estimates of net benefit for these models. Total sample size = 66,764 participants.

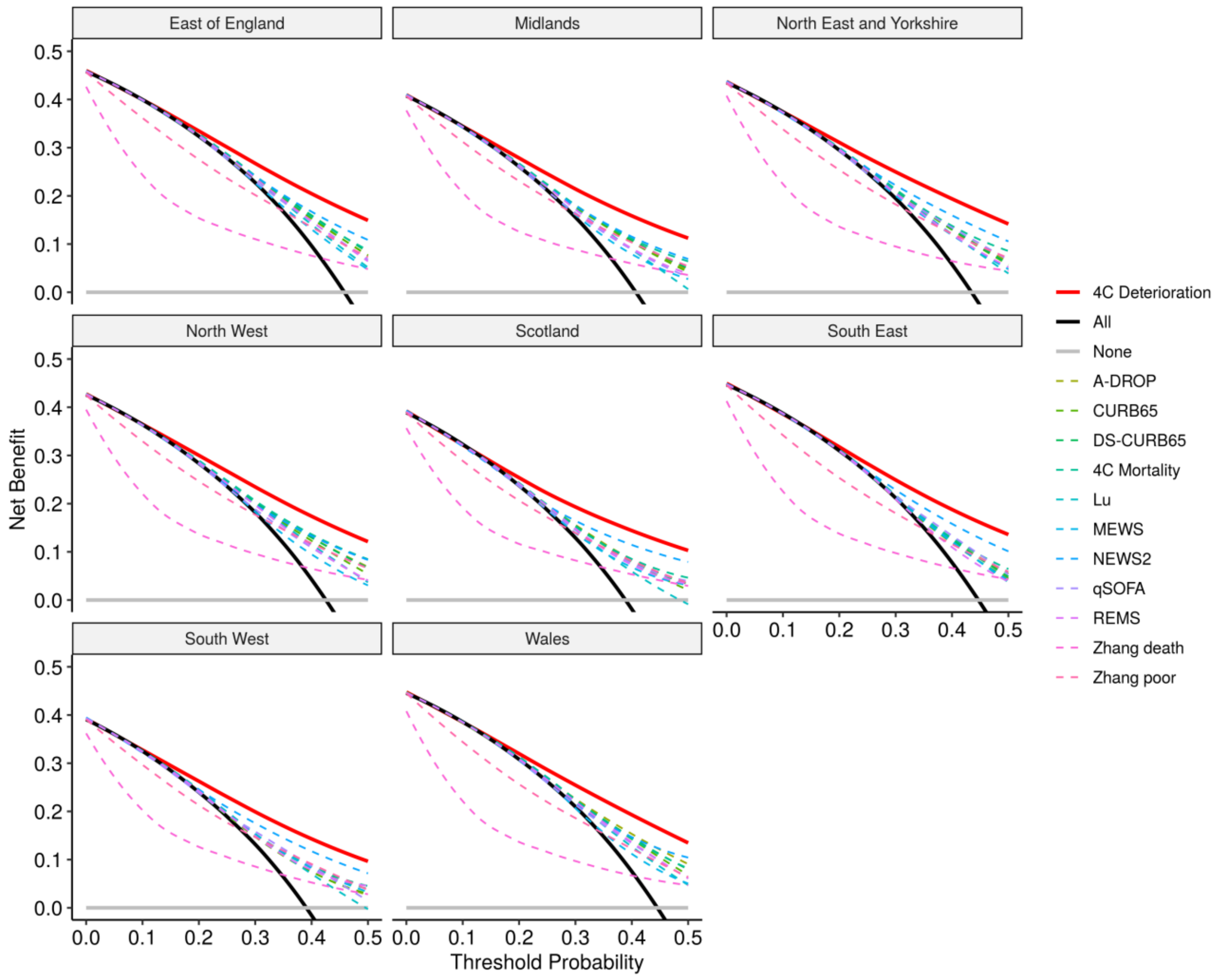

Supplementary Figure 5: Multivariable associations between predictors and outcome using alternative multiple imputation approach as sensitivity analysis.

Total development sample size = 66,764 participants. Black lines indicate point estimates; grey shaded regions indicate 95% confidence intervals.

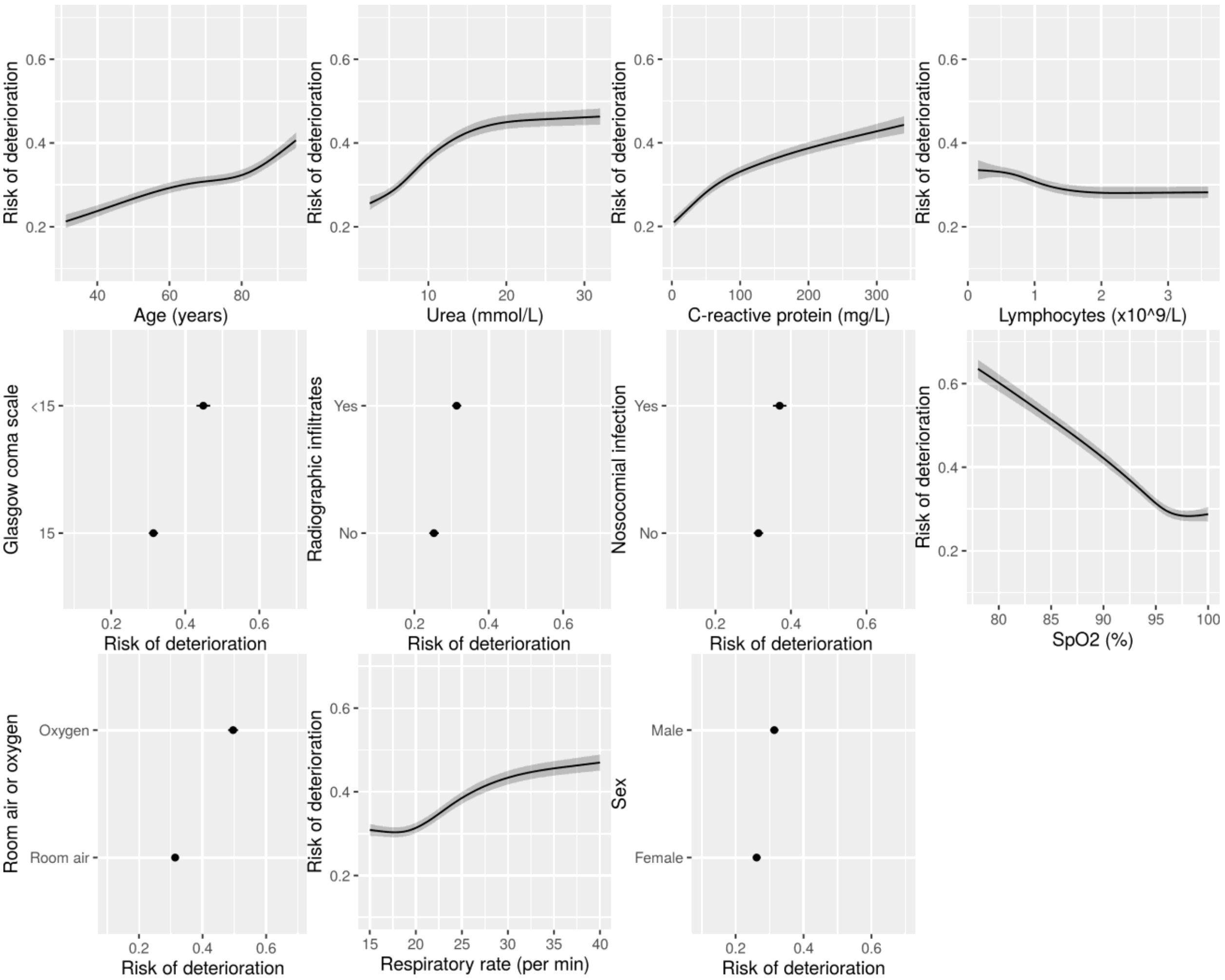

Supplementary Figure 6: Internal-external cross validation of model by NHS region using alternative multiple imputation approach as sensitivity analysis.

Pooled estimates are calculated through random-effects meta-analysis. Total sample size = 66,764 participants. Black squares indicate point estimates; bars indicate 95% confidence intervals; diamonds indicate pooled random-effects meta-analysis estimates.

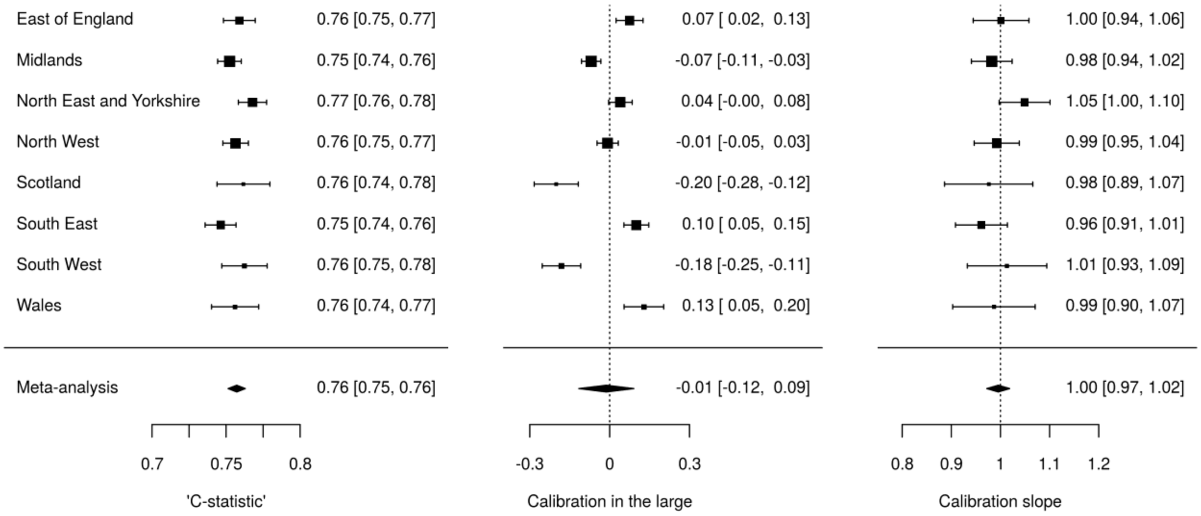

Supplementary Figure 7: Multivariable associations between predictors and outcome using alternative definition of nosocomial infection (>5 days from admission) as sensitivity analysis.

Total development sample size = 66,764 participants. Black lines indicate point estimates; grey shaded regions indicate 95% confidence intervals.

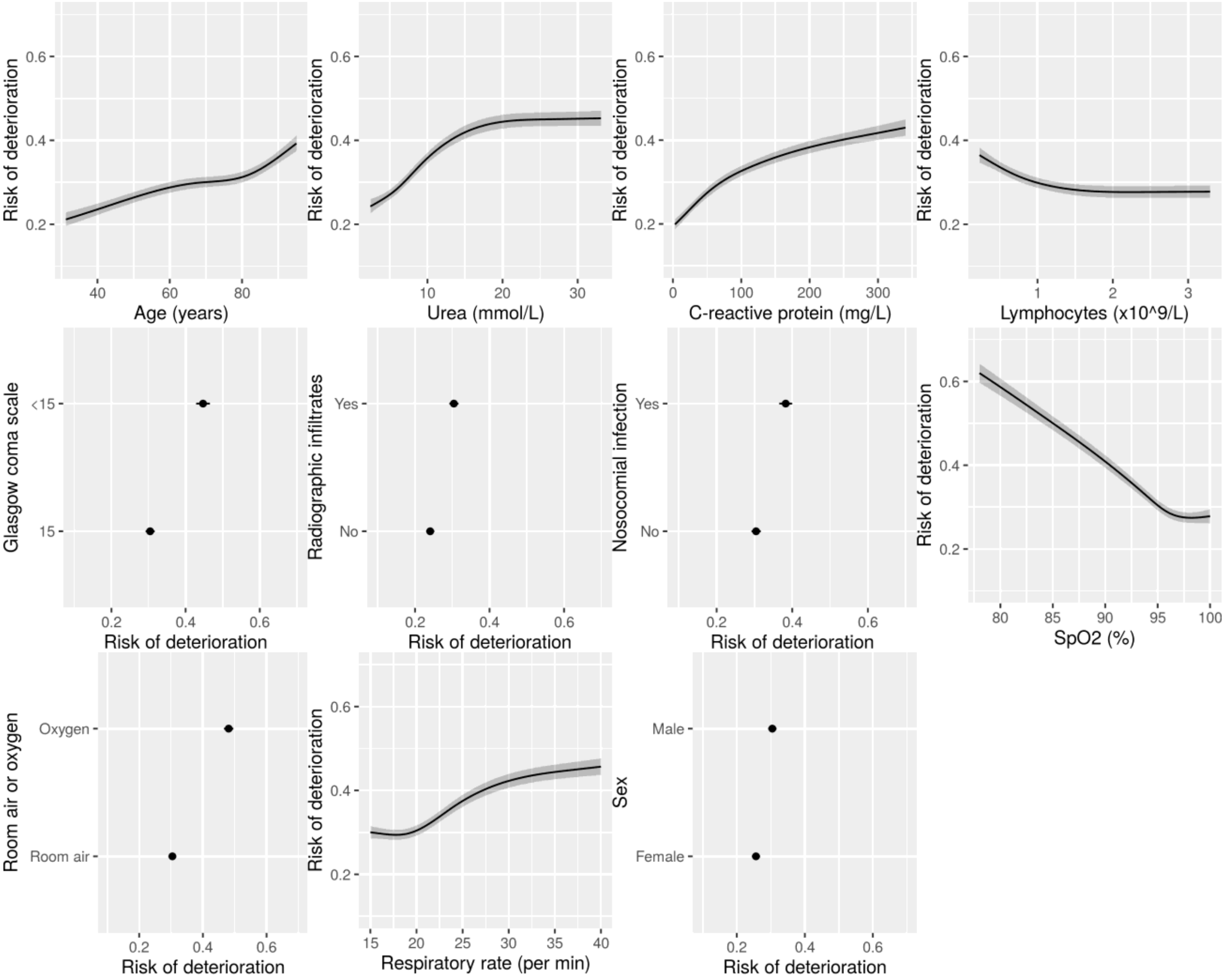

Supplementary Figure 8: Multivariable associations between predictors and outcome using alternative definition of nosocomial infection (>10 days from admission) as sensitivity analysis.

Total development sample size = 66,764 participants. Black lines indicate point estimates; grey shaded regions indicate 95% confidence intervals.

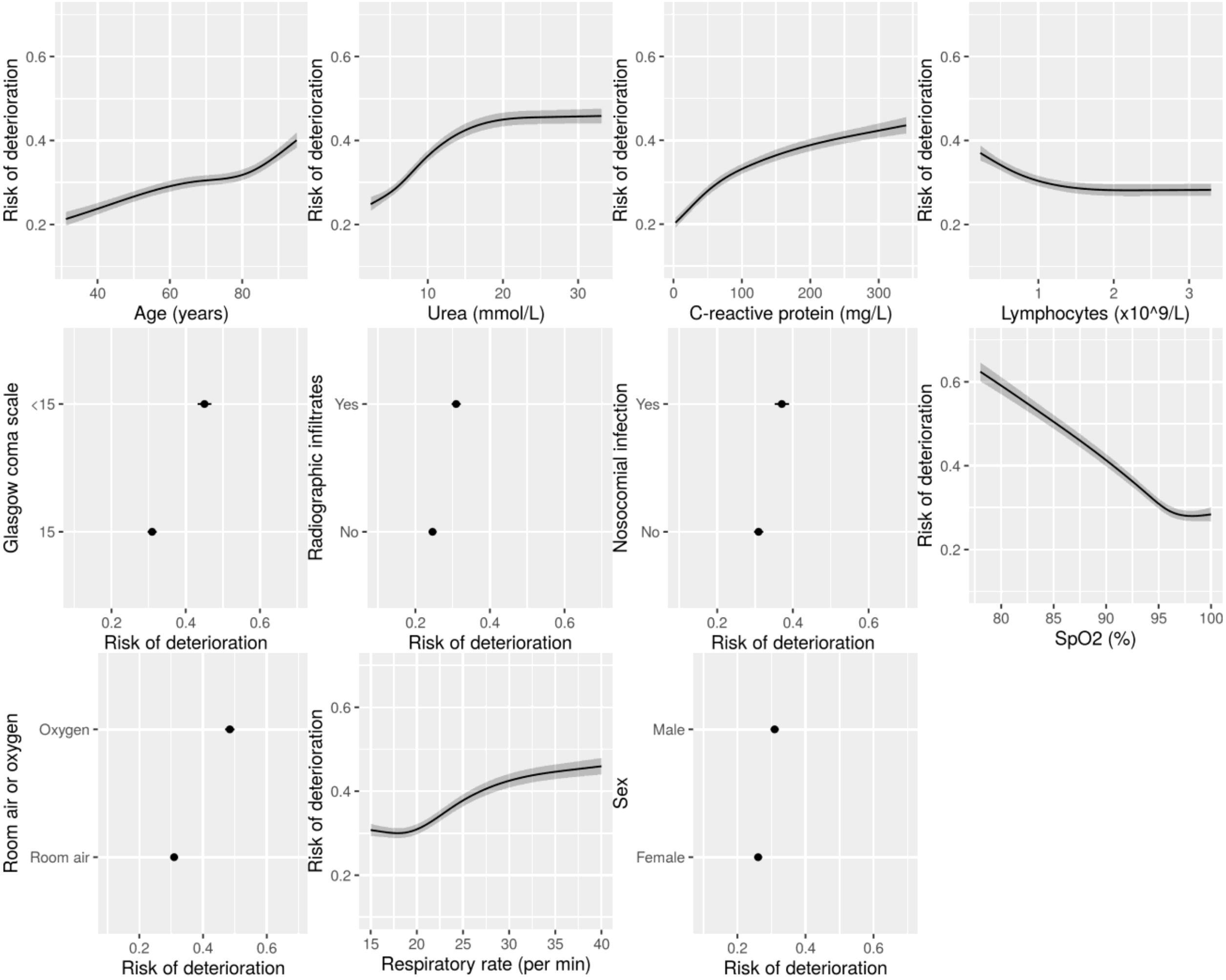

Supplementary Table 1: Characteristics of the study cohort, stratified by community vs nosocomial infection.

| Characteristic | Overall, N = 75,016 | No, N = 66,995[1] | Yes, N = 7,385[1] | (Missing), N = 636[1] |
|---|---|---|---|---|
| Age (years) | 75 (60, 84) | 74 (59, 84) | 80 (69, 87) | 78 (63, 87) |
| Sex | | | | |
| Female | 32,846 (44%) | 29,061 (43%) | 3,479 (47%) | 306 (48%) |
| Male | 42,026 (56%) | 37,815 (57%) | 3,882 (53%) | 329 (52%) |
| Unknown | 144 | 119 | 24 | 1 |
| Ethnicity | | | | |
| White | 55,079 (83%) | 48,719 (82%) | 5,900 (91%) | 460 (89%) |
| South Asian | 3,520 (5.3%) | 3,358 (5.6%) | 149 (2.3%) | 13 (2.5%) |
| Black | 2,554 (3.8%) | 2,432 (4.1%) | 107 (1.7%) | 15 (2.9%) |
| East Asian | 492 (0.7%) | 474 (0.8%) | 17 (0.3%) | 1 (0.2%) |
| Other | 4,848 (7.3%) | 4,514 (7.6%) | 307 (4.7%) | 27 (5.2%) |
| Unknown | 8,523 | 7,498 | 905 | 120 |
| Number of comorbidities | 1 (1, 2) | 1 (1, 2) | 2 (1, 3) | 1 (1, 2) |
| Unknown | 840 | 655 | 59 | 126 |
| Radiographic infiltrates | 29,594 (62%) | 28,460 (63%) | 1,047 (48%) | 87 (40%) |
| Unknown | 27,245 | 21,609 | 5,220 | 416 |
| Temperature (°C) | 37.2 (36.5, 38.1) | 37.2 (36.6, 38.1) | 36.8 (36.4, 37.6) | 36.7 (36.3, 37.1) |
| Unknown | 3,110 | 2,366 | 582 | 162 |
| Heart rate (per min) | 90 (78, 104) | 90 (78, 105) | 85 (72, 98) | 86 (74, 98) |
| Unknown | 3,389 | 2,535 | 685 | 169 |
| Respiratory rate (per min) | 20 (18, 26) | 22 (18, 26) | 18 (17, 20) | 18 (17, 20) |
| Unknown | 3,537 | 2,592 | 782 | 163 |
| Systolic blood pressure (mmHg) | 130 (114, 147) | 130 (114, 146) | 130 (113, 148) | 131 (115, 152) |
| Unknown | 3,192 | 2,359 | 674 | 159 |
| Diastolic blood pressure (mmHg) | 74 (64, 84) | 74 (65, 84) | 72 (62, 82) | 74 (65, 84) |
| Unknown | 3,335 | 2,485 | 690 | 160 |
| SpO2 (%) | 95 (92, 97) | 95 (92, 97) | 96 (94, 98) | 96 (95, 98) |
| Unknown | 3,760 | 2,873 | 724 | 163 |
| Room air or oxygen | | | | |
| Room air | 48,626 (69%) | 42,466 (67%) | 5,763 (88%) | 397 (87%) |
| Oxygen | 21,469 (31%) | 20,591 (33%) | 819 (12%) | 59 (13%) |
| Unknown | 4,921 | 3,938 | 803 | 180 |
| Glasgow coma scale | 15 (15, 15) | 15 (15, 15) | 15 (15, 15) | 15 (15, 15) |
| Unknown | 7,849 | 6,691 | 942 | 216 |
| Haemoglobin (g/L) | 128 (112, 142) | 129 (113, 143) | 111 (95, 128) | 120 (104, 135) |
| Unknown | 11,781 | 8,426 | 3,171 | 184 |
| White cell count (x10^9/L) | 7.5 (5.4, 10.7) | 7.6 (5.5, 10.7) | 7.2 (5.2, 10.2) | 8.4 (5.9, 11.3) |
| Unknown | 12,163 | 8,777 | 3,199 | 187 |
| Lymphocytes (x10^9/L) | 0.90 (0.60, 1.30) | 0.90 (0.60, 1.30) | 0.90 (0.60, 1.30) | 1.00 (0.60, 1.40) |
| Unknown | 12,377 | 8,953 | 3,236 | 188 |

| Characteristic | Overall, N = 75,016 | No, N = 66,995[1] | Yes, N = 7,385[1] | (Missing), N = 636[1] |
|---|---|---|---|---|
| Neutrophils (x10^9/L) | 5.8 (3.9, 8.7) | 5.8 (3.9, 8.7) | 5.3 (3.5, 8.2) | 6.4 (4.2, 9.4) |
| Unknown | 12,341 | 8,932 | 3,223 | 186 |
| Platelets (x10^9/L) | 221 (167, 291) | 219 (166, 287) | 246 (178, 332) | 241 (180, 312) |
| Unknown | 12,496 | 9,075 | 3,234 | 187 |
| Alanine aminotransferase (IU/L) | 25 (16, 43) | 25 (16, 43) | 21 (14, 37) | 20 (13, 39) |
| Unknown | 26,783 | 21,411 | 5,064 | 308 |
| Bilirubin (mg/dL) | 10 (7, 14) | 10 (7, 14) | 8 (5, 13) | 10 (7, 16) |
| Unknown | 22,979 | 17,790 | 4,883 | 306 |
| Urea (mmol/L) | 7 (5, 11) | 7 (5, 11) | 7 (5, 11) | 8 (5, 12) |
| Unknown | 18,554 | 14,652 | 3,661 | 241 |
| Creatinine (μmol/L) | 86 (67, 121) | 86 (67, 121) | 81 (60, 116) | 89 (67, 123) |
| Unknown | 12,693 | 9,286 | 3,214 | 193 |
| Sodium (mmol/L) | 137 (134, 140) | 137 (134, 140) | 137 (134, 141) | 137 (134, 141) |
| Unknown | 12,312 | 8,949 | 3,170 | 193 |
| C-reactive protein (mg/L) | 80 (33, 154) | 82 (34, 156) | 53 (20, 107) | 52 (12, 120) |
| Unknown | 16,353 | 12,558 | 3,561 | 234 |
| NHS region | | | | |
| East of England | 7,865 (10%) | 7,190 (11%) | 576 (7.8%) | 99 (16%) |
| London | 8,252 (11%) | 7,722 (12%) | 477 (6.5%) | 53 (8.3%) |
| Midlands | 15,599 (21%) | 14,059 (21%) | 1,464 (20%) | 76 (12%) |
| North East and Yorkshire | 10,310 (14%) | 9,393 (14%) | 862 (12%) | 55 (8.6%) |
| North West | 12,930 (17%) | 11,142 (17%) | 1,661 (22%) | 127 (20%) |
| Scotland | 3,068 (4.1%) | 2,812 (4.2%) | 243 (3.3%) | 13 (2.0%) |
| South East | 9,450 (13%) | 8,310 (12%) | 975 (13%) | 165 (26%) |
| South West | 3,919 (5.2%) | 3,495 (5.2%) | 407 (5.5%) | 17 (2.7%) |
| Wales | 3,623 (4.8%) | 2,872 (4.3%) | 720 (9.7%) | 31 (4.9%) |
| Deterioration | | | | |
| Ventilatory support or HDU/ICU | 15,089 (20%) | 14,435 (22%) | 592 (8.0%) | 62 (9.7%) |
| Died | 16,904 (23%) | 14,692 (22%) | 2,111 (29%) | 101 (16%) |
| No deterioration | 42,025 (56%) | 37,142 (55%) | 4,538 (61%) | 345 (54%) |
| (Missing) | 998 (1.3%) | 726 (1.1%) | 144 (1.9%) | 128 (20%) |
| Days from admission to COVID-19 assessment | 0 (0, 0) | 0 (0, 0) | 18 (11, 35) | 0 (0, 2) |
| Unknown | 1,775 | 1,611 | 104 | 60 |

[1]Statistics presented: median (IQR); n (%)

Supplementary Table 2: Final model parameters for 4C Deterioration prognostic model.

Parameters are pooled across multiply imputed datasets. Total development sample size = 66,764 participants.

| Characteristic | log(OR)[1] | 95% CI[1] | p-value |
|---|---|---|---|
| Intercept | 3.947 | 3.099, 4.569 | <0.001 |
| Age (years) | 0.0159 | 0.0132, 0.0206 | <0.001 |
| Age (spline 1) | -0.0132 | -0.0211, -0.0082 | <0.001 |
| Age (spline 2) | 0.1268 | 0.0889, 0.1833 | <0.001 |
| Sex | | | |
| Female | — | — | |
| Male | 0.2383 | 0.2057, 0.2770 | <0.001 |
| Nosocomial | | | |
| No | — | — | |
| Yes | 0.3026 | 0.2412, 0.3566 | <0.001 |
| Radiolographic infiltrates | | | |
| No | — | — | |
| Yes | 0.3185 | 0.2830, 0.3560 | <0.001 |
| Respiratory rate (per min) | -0.0145 | -0.0255, 0.0000 | 0.026 |
| Respiratory rate (spline 1) | 0.6048 | 0.4780, 0.7249 | <0.001 |
| Respiratory rate (spline 2) | -1.088 | -1.301, -0.8701 | <0.001 |
| SpO2 (%) | -0.0698 | -0.0772, -0.0620 | <0.001 |
| SpO2 (spline 1) | -0.0251 | -0.0378, -0.0083 | <0.001 |
| SpO2 (spline 2) | 1.010 | 0.5945, 1.263 | <0.001 |
| Room air or oxygen | | | |
| Room air | — | — | |
| Oxygen therapy | 0.7446 | 0.6952, 0.7729 | <0.001 |
| GCS | | | |
| 15 | — | — | |
| <15 | 0.6085 | 0.5456, 0.6479 | <0.001 |
| Urea (mmol/L) | 0.0533 | 0.0299, 0.0775 | <0.001 |
| Urea (spline 1) | 0.4277 | 0.1137, 0.7135 | 0.005 |
| Urea (spline 2) | -1.019 | -1.562, -0.4241 | <0.001 |
| C-reactive protein (mg/L) | 0.0098 | 0.0080, 0.0112 | <0.001 |
| C-reactive protein (spline 1) | -0.0389 | -0.0528, -0.0226 | <0.001 |
| C-reactive protein (spline 2) | 0.0577 | 0.0303, 0.0820 | <0.001 |
| Lymphocytes (x10^9/L) | -0.4708 | -0.6094, -0.2693 | <0.001 |
| Lymphocytes (spline 1) | 0.9161 | -0.3816, 1.841 | 0.11 |
| Lymphocytes (spline 2) | -1.235 | -3.237, 1.560 | 0.3 |

[1]OR = Odds Ratio, CI = Confidence Interval

Restricted cubic spline knot positions are:

- Age = 38.5, 67.7, 81.1, 92.9;

| Characteristic | log(OR)[1] | 95% CI[1] | p-value |
|---|---|---|---|

- Respiratory rate = 16, 19, 24, 37;
- SpO2 = 84, 94, 96, 100;
- Urea = 2.9, 5.7, 9.1, 25.3;
- C-reactive protein = 5, 45, 112, 294;
- Lymphocytes = 0.3, 0.7, 1.1, 2.4.

Supplementary Table 3: Validation in complete case London data as sensitivity analysis.

Models are shown for prediction of in-hospital clinical deterioration and are sorted by C-statistic. CITL = calibration-in-the-large. 'Original outcome' column indicates original intended outcome for each candidate model during development. CITL and slopes are not shown for points score models since they are not on probability scale.

| Score | Original outcome | n | C-statistic | CITL | Slope |
|---|---|---|---|---|---|
| 4C Deterioration | Deterioration (in-hospital) | 3769 | 0.76 (0.75 - 0.78) | -0.06 (-0.13 - 0.01) | 0.97 (0.89 - 1.04) |
| NEWS2 | Deterioration (1 day) | 6422 | 0.68 (0.67 - 0.7) | | |
| Zhang "death" | Mortality (in-hospital) | 6297 | 0.68 (0.67 - 0.69) | 2.33 (2.27 - 2.4) | 0.21 (0.18 - 0.23) |
| Zhang "poor" | Deterioration (in-hospital) | 6297 | 0.67 (0.66 - 0.69) | 0.63 (0.57 - 0.7) | 0.16 (0.14 - 0.18) |
| REMS | Mortality (in-hospital) | 6515 | 0.66 (0.65 - 0.67) | | |
| 4C Mortality | Mortality (in-hospital) | 5214 | 0.66 (0.64 - 0.67) | | |
| DS-CURB65 | Mortality (30 days) | 4832 | 0.66 (0.64 - 0.67) | | |
| A-DROP | Mortality (30 days) | 5051 | 0.64 (0.63 - 0.66) | | |
| CURB65 | Mortality (30 days) | 5080 | 0.64 (0.63 - 0.66) | | |
| qSOFA | Mortality (in-hospital) | 6754 | 0.63 (0.62 - 0.64) | | |
| MEWS | Deterioration (in-hospital) | 6575 | 0.62 (0.61 - 0.64) | | |
| Lu | Mortality (12 days) | 6627 | 0.61 (0.6 - 0.62) | | |

Supplementary Table 4: Validation in London data excluding deterioration events on day 0 as sensitivity analysis (n = 7,114 participants).

Models are shown for prediction of in-hospital clinical deterioration and are sorted by C-statistic. CITL = calibration-in-the-large. 'Original outcome' column indicates original intended outcome for each candidate model during development. CITL and slopes are not shown for points score models since they are not on probability scale.

| Score | Original outcome | C-statistic |
|---:|---:|---:|
| 4C Deterioration | Deterioration (in-hospital) | 0.75 (0.73 - 0.76) |
| 4C Mortality | Mortality (in-hospital) | 0.7 (0.68 - 0.71) |
| DS-CURB65 | Mortality (30 days) | 0.68 (0.66 - 0.69) |
| REMS | Mortality (in-hospital) | 0.67 (0.66 - 0.68) |
| A-DROP | Mortality (30 days) | 0.66 (0.65 - 0.68) |
| CURB65 | Mortality (30 days) | 0.66 (0.65 - 0.68) |
| NEWS2 | Deterioration (1 day) | 0.66 (0.65 - 0.68) |
| Zhang "poor" | Deterioration (in-hospital) | 0.66 (0.65 - 0.68) |
| Zhang "death" | Mortality (in-hospital) | 0.66 (0.64 - 0.67) |
| Lu | Mortality (12 days) | 0.64 (0.63 - 0.65) |
| qSOFA | Mortality (in-hospital) | 0.61 (0.6 - 0.63) |
| MEWS | Deterioration (in-hospital) | 0.6 (0.58 - 0.61) |

Supplementary Table 5: Validation in London data excluding patients still hospitalised at the end of follow-up as sensitivity analysis (n = 7,926 participants).

Models are shown for prediction of in-hospital clinical deterioration and are sorted by C-statistic. CITL = calibration-in-the-large. 'Original outcome' column indicates original intended outcome for each candidate model during development. CITL and slopes are not shown for points score models since they are not on probability scale.

| Score | Original outcome | C-statistic | CITL | Slope |
|---|---|---|---|---|
| 4C Deterioration | Deterioration (in-hospital) | 0.77 (0.76 - 0.78) | 0.1 (0.04 - 0.15) | 0.98 (0.92 - 1.04) |
| NEWS2 | Deterioration (1 day) | 0.69 (0.68 - 0.7) | | |
| 4C Mortality | Mortality (in-hospital) | 0.68 (0.66 - 0.69) | | |
| Zhang "poor" | Deterioration (in-hospital) | 0.68 (0.66 - 0.69) | 0.68 (0.61 - 0.74) | 0.17 (0.14 - 0.19) |
| Zhang "death" | Mortality (in-hospital) | 0.68 (0.66 - 0.69) | 2.37 (2.31 - 2.43) | 0.19 (0.16 - 0.22) |
| REMS | Mortality (in-hospital) | 0.67 (0.66 - 0.68) | | |
| DS-CURB65 | Mortality (30 days) | 0.67 (0.66 - 0.68) | | |
| CURB65 | Mortality (30 days) | 0.66 (0.65 - 0.67) | | |
| A-DROP | Mortality (30 days) | 0.66 (0.64 - 0.67) | | |
| qSOFA | Mortality (in-hospital) | 0.63 (0.62 - 0.64) | | |
| MEWS | Deterioration (in-hospital) | 0.63 (0.61 - 0.64) | | |
| Lu | Mortality (12 days) | 0.62 (0.61 - 0.63) | | |

Supplementary Table 6: Validation in London data stratified by (a) community (n = 7,771 participants) vs (b) nosocomial infection (n = 481 participants) as sensitivity analysis.

Models are shown for prediction of in-hospital clinical deterioration and are sorted by C-statistic. CITL = calibration-in-the-large. 'Original outcome' column indicates original intended outcome for each candidate model during development. CITL and slopes are not shown for points score models since they are not on probability scale.

(a)

| Score | Original outcome | C-statistic | CITL | Slope |
|---:|---:|---:|---:|---:|
| 4C Deterioration | Deterioration (in-hospital) | 0.77 (0.76 - 0.78) | -0.01 (-0.07 - 0.04) | 0.97 (0.91 - 1.03) |
| NEWS2 | Deterioration (1 day) | 0.7 (0.68 - 0.71) | | |
| Zhang "death" | Mortality (in-hospital) | 0.67 (0.66 - 0.69) | 2.21 (2.15 - 2.28) | 0.19 (0.16 - 0.22) |
| 4C Mortality | Mortality (in-hospital) | 0.67 (0.66 - 0.68) | | |
| REMS | Mortality (in-hospital) | 0.67 (0.66 - 0.68) | | |
| Zhang "poor" | Deterioration (in-hospital) | 0.67 (0.66 - 0.68) | 0.51 (0.45 - 0.58) | 0.16 (0.13 - 0.18) |
| DS-CURB65 | Mortality (30 days) | 0.66 (0.65 - 0.67) | | |
| A-DROP | Mortality (30 days) | 0.65 (0.64 - 0.66) | | |
| CURB65 | Mortality (30 days) | 0.65 (0.64 - 0.66) | | |
| MEWS | Deterioration (in-hospital) | 0.63 (0.62 - 0.65) | | |
| qSOFA | Mortality (in-hospital) | 0.63 (0.62 - 0.64) | | |
| Lu | Mortality (12 days) | 0.62 (0.6 - 0.63) | | |

(b)

| Score | Original outcome | C-statistic | CITL | Slope |
|---:|---:|---:|---:|---:|
| 4C Deterioration | Deterioration (in-hospital) | 0.72 (0.67 - 0.77) | 0.37 (0.17 - 0.56) | 0.94 (0.68 - 1.2) |
| 4C Mortality | Mortality (in-hospital) | 0.67 (0.62 - 0.72) | | |
| Zhang "poor" | Deterioration (in-hospital) | 0.65 (0.6 - 0.71) | 1 (0.75 - 1.26) | 0.1 (0.01 - 0.19) |
| Zhang "death" | Mortality (in-hospital) | 0.65 (0.6 - 0.71) | 2.95 (2.67 - 3.22) | 0.12 (0.01 - 0.23) |
| DS-CURB65 | Mortality (30 days) | 0.65 (0.6 - 0.7) | | |
| NEWS2 | Deterioration (1 day) | 0.65 (0.6 - 0.7) | | |
| Lu | Mortality (12 days) | 0.64 (0.6 - 0.69) | | |
| A-DROP | Mortality (30 days) | 0.64 (0.59 - 0.69) | | |
| CURB65 | Mortality (30 days) | 0.63 (0.58 - 0.68) | | |
| MEWS | Deterioration (in-hospital) | 0.6 (0.55 - 0.65) | | |
| REMS | Mortality (in-hospital) | 0.6 (0.55 - 0.65) | | |
| qSOFA | Mortality (in-hospital) | 0.59 (0.54 - 0.63) | | |

Supplementary Table 7: Validation parameters of prognostic model among community-acquired cases in London, excluding those with symptom onset recorded after admission date, as sensitivity analysis (n = 7,167 participants).

Models are shown for prediction of in-hospital clinical deterioration and are sorted by C-statistic. CITL = calibration-in-the-large. 'Original outcome' column indicates original intended outcome for each candidate model during development. CITL and slopes are not shown for points score models since they are not on probability scale.

| Score | Original outcome | C-statistic | CITL | Slope |
|---:|---:|---:|---:|---:|
| 4C Deterioration | Deterioration (in-hospital) | 0.77 (0.75 - 0.78) | -0.01 (-0.07 - 0.04) | 0.97 (0.9 - 1.03) |
| NEWS2 | Deterioration (1 day) | 0.69 (0.68 - 0.71) | | |
| Zhang "death" | Mortality (in-hospital) | 0.67 (0.66 - 0.69) | 2.24 (2.17 - 2.31) | 0.19 (0.15 - 0.22) |
| REMS | Mortality (in-hospital) | 0.67 (0.65 - 0.68) | | |
| Zhang "poor" | Deterioration (in-hospital) | 0.67 (0.65 - 0.68) | 0.56 (0.5 - 0.63) | 0.15 (0.12 - 0.18) |
| 4C Mortality | Mortality (in-hospital) | 0.66 (0.65 - 0.68) | | |
| DS-CURB65 | Mortality (30 days) | 0.66 (0.65 - 0.67) | | |
| CURB65 | Mortality (30 days) | 0.65 (0.64 - 0.66) | | |
| A-DROP | Mortality (30 days) | 0.65 (0.63 - 0.66) | | |
| qSOFA | Mortality (in-hospital) | 0.63 (0.62 - 0.64) | | |
| MEWS | Deterioration (in-hospital) | 0.63 (0.61 - 0.64) | | |
| Lu | Mortality (12 days) | 0.61 (0.6 - 0.62) | | |

Supplementary Table 8: Validation parameters of prognostic model in London cohort using alternative multiple imputation approach as sensitivity analysis (n = 8,252 participants).

Models are shown for prediction of in-hospital clinical deterioration and are sorted by C-statistic. CITL = calibration-in-the-large. 'Original outcome' column indicates original intended outcome for each candidate model during development. CITL and slopes are not shown for points score models since they are not on probability scale.

| Score | Original outcome | C-statistic | CITL | Slope |
|---|---|---|---|---|
| 4C Deterioration | Deterioration (in-hospital) | 0.76 (0.75 - 0.77) | 0 (-0.05 - 0.05) | 0.93 (0.88 - 0.99) |
| NEWS2 | Deterioration (1 day) | 0.68 (0.67 - 0.7) | | |
| 4C Mortality | Mortality (in-hospital) | 0.66 (0.65 - 0.67) | | |
| REMS | Mortality (in-hospital) | 0.66 (0.65 - 0.67) | | |
| DS-CURB65 | Mortality (30 days) | 0.65 (0.64 - 0.66) | | |
| Zhang "death" | Mortality (in-hospital) | 0.65 (0.64 - 0.66) | 2.07 (2 - 2.13) | 0.14 (0.11 - 0.16) |
| Zhang "poor" | Deterioration (in-hospital) | 0.65 (0.63 - 0.66) | 0.33 (0.26 - 0.4) | 0.11 (0.09 - 0.13) |
| CURB65 | Mortality (30 days) | 0.64 (0.63 - 0.66) | | |
| A-DROP | Mortality (30 days) | 0.64 (0.63 - 0.65) | | |
| qSOFA | Mortality (in-hospital) | 0.62 (0.61 - 0.64) | | |
| MEWS | Deterioration (in-hospital) | 0.62 (0.61 - 0.63) | | |
| Lu | Mortality (12 days) | 0.61 (0.6 - 0.62) | | |
